# Explaining the counter-intuitive effectiveness of trophectoderm biopsy for PGT-A using computational modelling

**DOI:** 10.1101/2023.12.12.23299850

**Authors:** Benjamin M Skinner, Manuel Viotti, International Registry of Mosaic Embryo Transfers (IRMET), Darren K Griffin, Peter JI Ellis

## Abstract

Preimplantation genetic testing for aneuploidy (PGT-A) is one of the most controversial topics in reproductive medicine, with disagreements over the apparently contradictory results of randomised controlled trials, non-selection trials and outcome data analyses. Data from live birth outcomes largely suggest that fully euploid biopsies are associated with positive live birth rates, while fully aneuploid biopsies are not. However, the possible confounding effects of chromosomal mosaicism (when either the whole embryo, the biopsy result (or both) contain an admixture of euploid and aneuploid cells) is frequently cited as a reason why PGT-A should not be performed. Previous computer models have indicated that a mosaic result is a poor indicator of the level of mosaicism of the rest of the embryo, and it is thus unwise to use mosaic PGT-A results when selecting embryos for transfer. Here we developed a computational model, *tessera*, to create virtual embryos for biopsy, allowing us to vary the number of cells in the simulated embryo and biopsy, the proportion of aneuploid cells and the degree of juxtaposition of those cells. Analysis of approximately 1 million virtual embryos showed that “100% euploid” and “100% aneuploid” biopsy results are relatively accurate predictors of the remainder of the embryo, while mosaic biopsy results are poor predictors of the proportion of euploid and aneuploid cells in the rest of the embryo. Within mosaic embryos, ‘clumping’ of aneuploid cells further reduces the accuracy of biopsies in assaying the true aneuploidy level of any given embryo. Nonetheless - and somewhat counterintuitively - biopsy results can still be used with some confidence to rank embryos within a cohort. Our simulations help resolve the apparent paradox surrounding PGT-A: the biopsy result is poorly predictive of the absolute level of mosaicism of a single embryo, but may be applicable nonetheless in making clinical decisions on which embryos to transfer.

## Introduction

Preimplantation Genetic Testing for Aneuploidy (PGT-A) is one of the most controversial areas of treatment in reproductive medicine. Since its inception, questions about its efficacy have led to entrenched points of view both in favour and against its use (Griffin and Ogur, 2018; Victor et al., 2020). While randomised controlled trials (e.g. Yan et al., 2021), mostly in good prognosis patients, often point to minimal or no efficacy (at least in younger women), non-selection trials (e.g. Tiegs et al., 2021; Wang et al., 2021; Yang et al., 2021) provide strong evidence that embryos diagnosed as fully aneuploid rarely lead to live births and frequently miscarry. Some consensus can be arrived at along the lines of a) PGT-A does not improve cumulative pregnancy/live birth rates (some would say it was never designed to), but b) PGT-A does nonetheless improve pregnancy/live birth rates per embryo transfer (though some would say that this is not an accurate measure of efficacy). Key points of debate and disagreement still persist however as follows: First, the extent to which PGT-A improves pregnancy/live birth rate per cycle. While raw outcome data points to a clear positive benefit of PGT-A (Sanders et al., 2021), this work has been challenged by Roberts et al., (2022) as “naïve analysis”; they performed a more in-depth logistic regression including confounders such as patient history, treatment characteristics and year of treatment, and suggested that PGT-A may be detrimental overall. Second, the prospect that the process of biopsying the embryo could lead to damage and therefore impaired developmental potential. While Scott et al., (2013) have provided compelling evidence that this is not the case in their own setting, worldwide roll-out could theoretically lead to suboptimal practices in some clinics. Third, the possible confounding effects of euploid/aneuploid mosaicism. Specifically, the likely role of embryo “self-correction” (high levels of embryonic mosaicism earlier in development demonstrably reduce in later stages), and, pertinent to this study, whether any mosaic result found in the biopsy accurately reflects that of the rest of the embryo.

Currently, PGT-A involves the removal of a 5-10 cell biopsy from the trophectoderm (TE) of a ∼150-200 cell blastocyst embryo. Most PGT-A performed today entails assessment of the biopsy’s ploidy by low-read whole genome sequencing and subsequent read depth counting (reviewed in Viotti, 2020). The end result is a profile with wide dynamic range that is well established, with considerable accuracy, to assay for the proportion of euploid and aneuploid cells in known admixtures of five or more cells. The subsequent translation to a five-cell embryo biopsy is inferred with some confidence therefore and, by definition, if mosaicism is present in the biopsy, the embryo as a whole was mosaic prior to biopsy. The *level* of mosaicism (i.e. the proportion of aneuploid to euploid cells) among the un-biopsied remainder of the embryo however is not reliably inferred. That is, if two out of the five cells in the biopsy are aneuploid, this does not necessarily mean that the remainder of the embryo is similarly 40% aneuploid and 60% euploid. There are three reasons for this: First, from basic probabilities of sampling from a mixed population, a five-cell biopsy will not always reflect this ratio. Second, the majority of euploid/aneuploid mosaics arise by post-zygotic chromosomal segregation errors; populations of aneuploid cells are thus clonal and would be expected to be in rough juxtaposition to one another. In other words, a mosaic embryo would be expected to have “clumps” of euploid and aneuploid cells, non-homogeneously distributed. Third, there is evidence that the TE and ICM (inner cell mass) can have different levels of aneuploidy or mosaicism (Griffin et al., 2022; Ren et al., 2022). The blastocyst thus either actively expels aneuploid cells to the trophectoderm and other structures (blastocoel, surrounding degenerate cells) or the ICM disproportionately disfavours the growth of aneuploid cells (Griffin et al., 2022). Indeed, as more data emerges, it appears that the majority of embryos from both healthy and infertile couples are mosaic to some degree (Coticchio et al., 2021; Griffin et al., 2022). Moreover, human embryonic development is characterised, chromosomally, to be complex, fluid and dynamic, with demonstrably fewer chromosome abnormalities present at day 3 compared to day 5 (Coticchio et al., 2021; Harton et al., 2017).

Experimental data provides strong evidence that, for the most part, the biopsy result obtained accurately represents the chromosome constitution of the rest of the embryo (Kim et al., 2022; Navratil et al., 2020; Victor et al., 2019). The majority of biopsy results are however “100% euploid” or “100% aneuploid”, with mosaic results comprising ∼4-20% of returns. Furthermore, 100% euploid diagnoses may not have detected low level euploid/aneuploid mosaicism by virtue of the fact that, simply, no aneuploid cells were biopsied. Initial reports of live births following mosaic embryo diagnoses (Greco et al., 2015) were followed by the establishment of a registry logging the transfers of mosaic embryos and their clinical outcomes (Viotti et al., 2023, 2021). Data shows that live birth rate per embryo transfer is generally lower in mosaic embryos than embryos with a “100% euploid result”, but that other health outcomes for babies born from the mosaic or euploid groups seem largely indistinguishable (Viotti et al., 2023, 2021).

Computational models provide a useful adjunct to clinical outcome data studies, allowing multiple iterations of the same biological scenario to be tested. In this study, we explored, comprehensively, the utility of embryo biopsy using computational modelling. Previous modelling of embryo biopsy as a sampling approach using a hypergeometric distribution model demonstrated limitations in how much information can be obtained from a single biopsy (Gleicher et al., 2017). This modelling assumed aneuploid cells were randomly distributed within an embryo; however, their clonal origin suggests this to be unlikely (Mantikou et al., 2012). Clustered aneuploid cells in a biopsy do not fit a hypergeometric distribution and so the previous model could have *overstated* the utility of biopsying. If we were to ask the question “does a mosaic diagnosis accurately predict the level of mosaicism in the rest of the embryo?” we need to define what we mean by “accurately” – within 1%, 5%, 10% or 20% for instance. Finally, even if we cannot accurately predict the absolute level of embryo mosaicism, does this mean that the *ranking* of embryos based on the level of mosaicism seen is also invalid?

With the above in mind, we developed a more extensive computational model of embryo biopsy, allowing us to explore the scenarios in which biopsies do, or do not, provide useful information. We were interested not only in the proportion of aneuploid cells in both the biopsy and the whole embryo, but also how dispersed or clustered those cells are, as well as the size of the biopsy. We related this data back to previously described classifications of aneuploidy rates into low and high-level mosaicism (Munné et al., 2020) and in the PGDIS position statement on the transfer of mosaic embryos (Leigh et al., 2022). This allowed us to resolve the paradoxical utility of trophectoderm biopsy for PGT-A via a simple maxim: *although the information contained in the biopsy is highly imperfect, even imperfect information is clinically useful*.

## Methods

In order to develop a “virtual embryo” computational model, we used R (R Core Team, 2022). The virtual embryo was implemented in the *tessera* R package, available from https://github.com/bmskinner/tessera and allowed us to design virtual embryos with specific proportions of aneuploid cells, dispersals of those cells and to take different sizes of biopsy in multiple iterations.

The embryo was implemented as a Fibonacci lattice: projection of a Fibonacci spiral into spherical coordinates, providing a sphere with evenly spaced points of desired number at the surface (González, 2009; Swinbank and Purser, 2006). This represents the TE (for simplicity, we do not consider the ICM in this model). Each cell in the embryo can be euploid or aneuploid; when an embryo is created using this model, the proportion of aneuploid cells is specified, and the level to which the aneuploid cells are dispersed on a scale of 0-1. The overall placement strategy models a biological situation in which a variable number of “seed” progenitor cells undergo mal-segregation at an early-stage embryonic development, followed by clonal expansion of these aneuploid cells within their immediate vicinity. The “dispersion” parameter controls the number of initial seed regions, ranging from 0 (no dispersion, one single clump of aneuploid cells is present) to 1 (maximal dispersion, each aneuploid cell in the embryo is placed non-adjacent to other aneuploid cells if possible).

Technically, the dispersion is set by seeding initial aneuploid cells that are not adjacent to another aneuploid cell, and which can grow into aneuploid patches. As dispersion increases, so does the number of initial seeds. When all seeds have been placed, or no more seeds can be placed without being adjacent to an existing aneuploid cell, any remaining aneuploid cells required are selected randomly from the euploid cells adjacent to at least one aneuploid cell. The placement of aneuploid cells is random within these constraints, controlled by R’s random number generator. An overall seed for the generator can be set for each embryo to allow reproducibility.

We then repeatedly biopsied the embryo to yield all possible biopsies from that embryo. A biopsy of desired size *n* is taken by sampling a cell and its *n-1* closest neighbours. This is repeated for each cell in the embryo. The number of aneuploid cells in each biopsy is then counted and aggregated.

We considered four parameters: aneuploidy, dispersal, embryo size and biopsy size. We simulated embryos with aneuploidy from 0% to 100% in 1% intervals, and dispersal of aneuploid cells, from 0 (clustered) to 1 (fully dispersed) in 0.01fractional intervals. Four embryo sizes were modelled: 100, 150, 200 and 250 cells, and twelve biopsy sizes: 3, 4, 5, 6, 7, 8, 9, 10, 15, 20, 25 and 30 cells. For each combination of aneuploidy, dispersal, embryo size and biopsy size we generated 100 embryos using different random number seeds.

Given the large amount of data, unless otherwise stated, the results presented herein use a biopsy size of 5 cells and embryo size of 200 cells, as aneuploidy and dispersal were more important than embryo or biopsy size to the results. Simulated data were created using tessera v0.6 and results were visualised using ggplot2 (Wickham, 2016). The tessera package also contains a visualisation tool in Shiny (Chang et al., 2022) for exploring the impacts of aneuploidy and dispersal on biopsy results. Tessera can be run in a local web browser, and is also available for basic visualisation via web server at https://reproduction.essex.ac.uk.

### Statistical analysis of clinical data

We assessed the latest clinical data for empirical support for our model outputs. Data were obtained from 1733 embryos by the International Registry of Mosaic Embryo Transfers (IRMET), of which 1000 have previously been reported in Viotti et al. (2021), and 733 collected since. The analyses presented here were approved by the IRB of the Zouves Foundation (OHRP IRB00011505, Protocol #0002). Embryo were evaluated for chromosomal abnormalities in trophectoderm biopsies by PGT-A (Veisieq, Vitrolife) and classified as ‘mosaic’ if the results indicated intermediate copy number for any genomic region within the assay’s resolution (>20Mb) in the 20-80% interval between whole chromosome numbers, as laid out by the position statement issued by the Preimplantation Genetic Diagnosis International Society (PGDIS) (Leigh et al., 2022). Embryos were classified following Viotti et al. (2021) as either whole chromosome mosaics (at least one whole chromosomal aneuploidy and zero or more segmental aneuploidies) or segmental mosaics (at least one segmental aneuploidy and no whole chromosomal aneuploidy).

Statistical analysis was performed in R. Data were analysed by logistic regression, modelling successful outcome as dependent on the type of aneuploidy and the level of aneuploidy. Comparisons between categorical groups were performed with two-tailed chi-square tests with Bonferroni correction for multiple testing.

Scripts used to generate all analyses and figures presented are available at https://github.com/bmskinner/embryo_biopsy.

## Results

We developed the R package ‘tesser’ to simulate trophectoderm biopsies, allowing us to compare the effects of changing embryo parameters on biopsy results. Figure 1 shows two example embryos, both with 20% aneuploidy, but different dispersal of the aneuploid cells (Fig 1 A,B). The distribution of biopsies obtained from these embryos consequently also differs.

**Figure 1:**
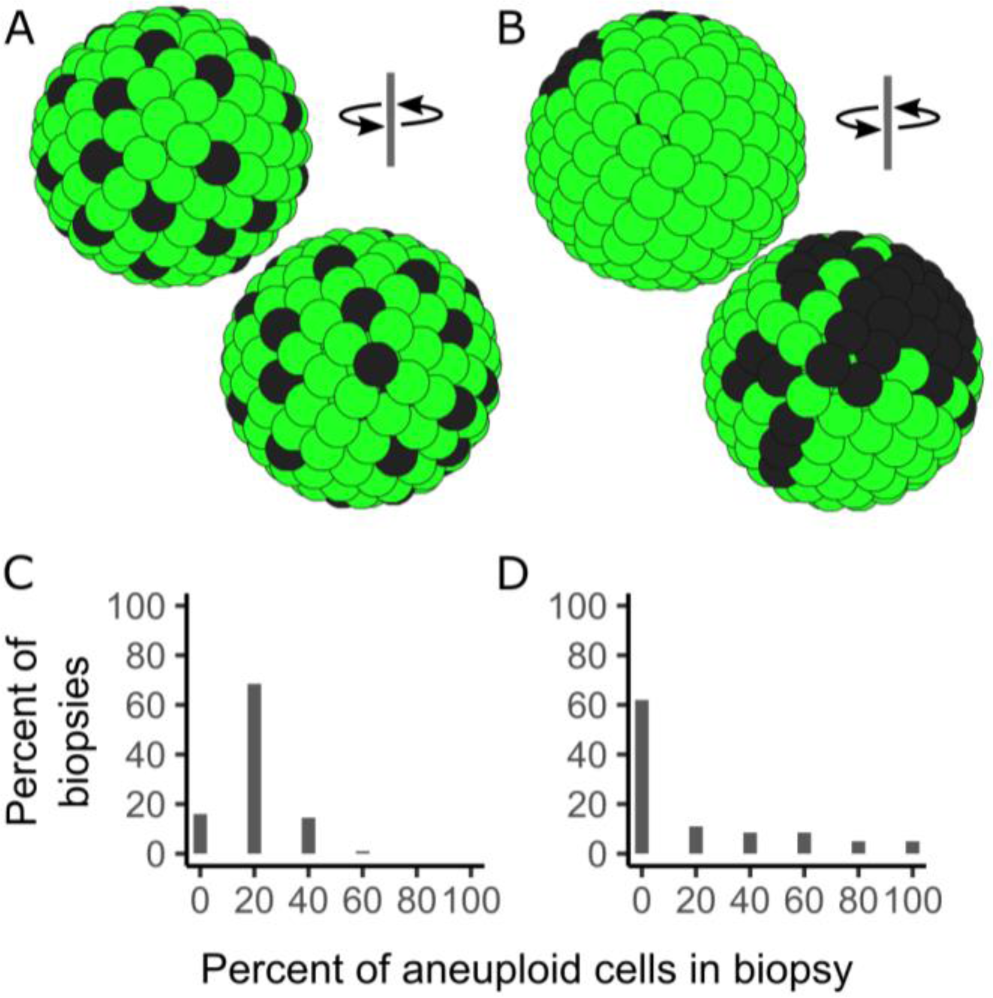
Model embryos created using tessera with 20% aneuploid cells (dark grey) and 80% euploid cells (light green), shown from front and back. Embryos have either high (A) or low (B) dispersal of the aneuploid cells. The potential biopsies obtained from each differ (graph C for embryo A, graph D for embryo B). For the clustered embryo (B) most biopsies are not representative of the mosaicism level present in the embryo (D); the reverse is true for the dispersed embryo.

In the dispersed embryo (Fig 1A), the majority of biopsies also have 20% aneuploidy (1 aneuploid cell in the biopsy; Fig 1C), but in clustered embryo (Fig 1B) the majority of the biopsies come from the euploid region of the embryo and have no aneuploid cells (Fig 1D). Biopsies taken from a clustered embryo are hence less likely to be representative of the embryo than biopsies taken from the dispersed embryo.

We used this approach to explore the impact of aneuploidy level, dispersal, biopsy size and embryo size by simulating and biopsying multiple randomly generated embryos for each parameter combination.

### Aneuploidy level and dispersal both affect biopsy accuracy

In the first step, we kept dispersal high, and varied the level of aneuploidy. We visualised the overall accuracy of the biopsies by comparing the biopsy aneuploidy level to the overall embryo aneuploidy level (Figure 2A). We saw an increase in the error as aneuploidy increases towards 50%. This matches expectations: the more cells of a single type are present in the embryo, the more likely a biopsy is to reflect those cells. There is also a decrease in error as aneuploidy increases from 50% to 100% and the cells again become more consistent.

**Figure 2:**
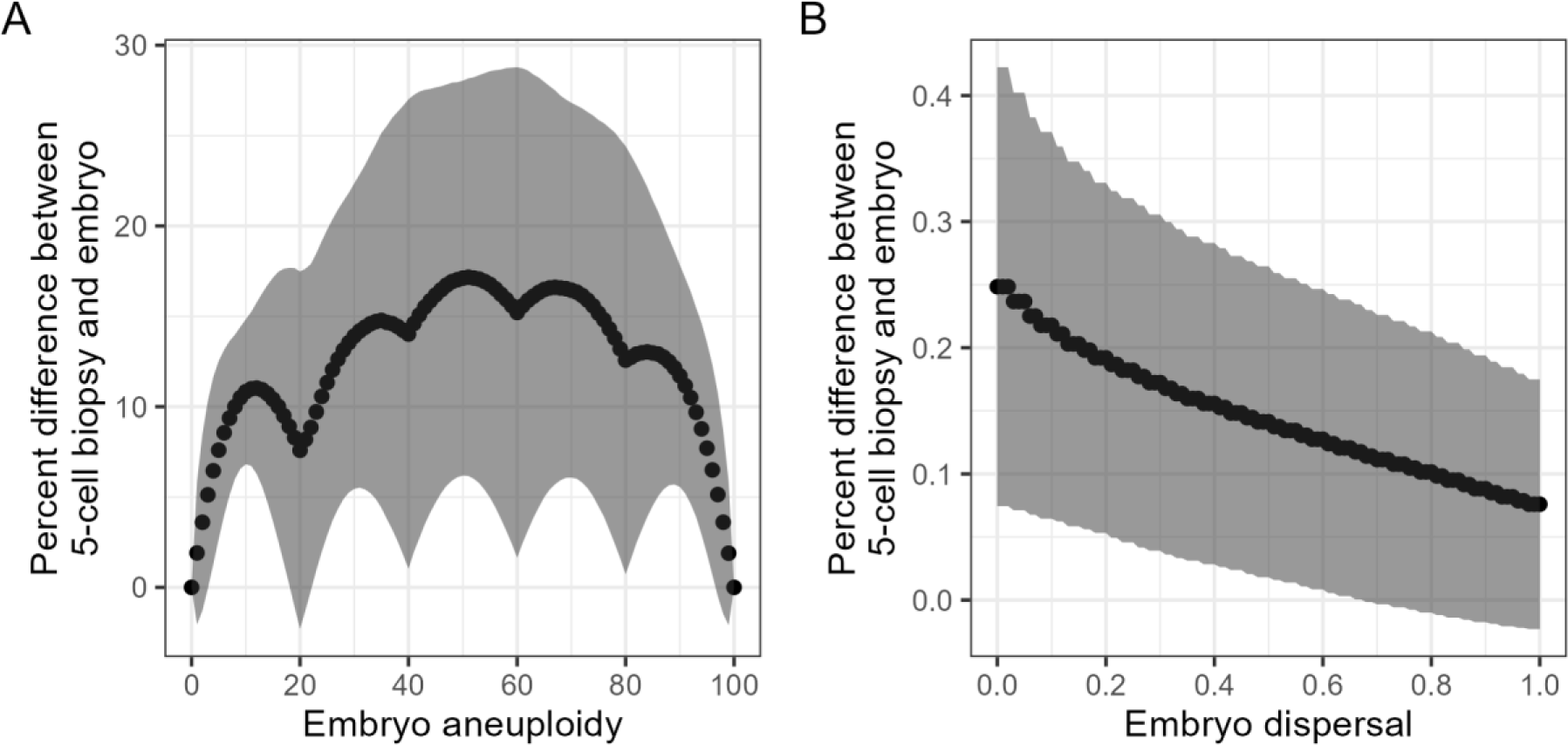
Difference between the aneuploidy of a 5-cell biopsy and the aneuploidy of the embryo for embryos with increasing levels of (A) aneuploidy at constant dispersal of 1 and (B) increasing dispersal with constant aneuploidy of 20%. Values show the mean and standard deviation of 100 replicates. Error is highest at 40%-60% aneuploidy, and at low dispersals. The local minima in the aneuploidy chart are due to the 5-cell biopsy size, allowing exact matches to the biopsy at these aneuploidies.

If aneuploidy is held constant at 20%, and dispersal of aneuploid cells is varied, we obtain the pattern in Fig 2B: greatest error at low dispersal, decreasing as dispersal increases. This also matches expectations: a biopsy from a highly dispersed embryo will be more likely to reflect the embryo than a biopsy taken from a patch of clustered aneuploid cells.

### Hard classification thresholds cause lower accuracy in classifying embryos close to the boundary

The difference between the level of mosaicism in the biopsy and that in the embryo is one useful measure, but there are other classification methods used in clinics. The PGDIS (Preimplantation Genetic Diagnosis International Society) described classification levels for aneuploidy that are followed or adapted by many fertility clinics (Leigh et al., 2022; Munné et al., 2020), hereafter termed ‘embryo classes’ for brevity. Under the position statement, the following classification system is possible: 0-<20% aneuploidy is considered ‘euploid’, 20-<50% ‘low level’ mosaic, 50-80% ‘high level’ mosaic, and >80-100% ‘aneuploid’. We used these classifications as a more practical assessment of biopsy accuracy: what percentage of biopsies from an embryo are in the same class as the embryo itself?

Figure 3 shows this embryo classification accuracy for dispersal and aneuploidy combinations as a heatmap. Each combination is calculated as the mean from 100 embryos. Large differences are seen between the accuracies of biopsies from embryos in the different embryo classes become apparent. Generally, high dispersal of aneuploid cells is more accurate than clustered cells, and extremes of aneuploidy are more accurate than intermediate mosaic levels of aneuploidy. Overall classification accuracy is lowest in the low-level mosaic class and at the boundaries between classes.

**Figure 3:**
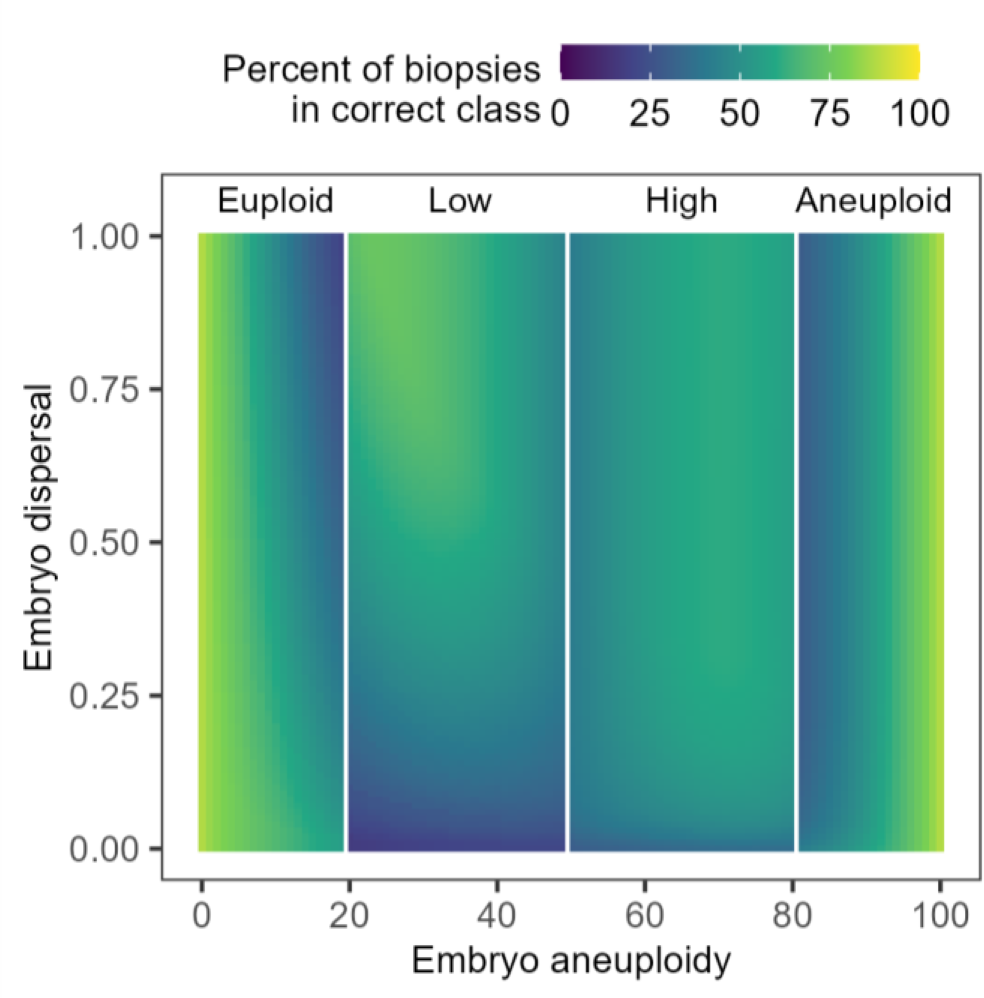
The percentage of biopsies in the same class as the embryo from which they came, for all combinations of aneuploidy and dispersal. Each combination represents the mean of 100 embryos. Accuracy is lower in embryos with low dispersal, especially in ‘low-level’ mosaic embryos.

### Increasing biopsy size does not greatly increase accuracy in clustered embryos

All models presented so far have used a biopsy size of 5 cells. In PGT-A biopsies tend to be 5-10 cells, so we tested the effect of varying biopsy size from 3 cells to an unrealistically high 30 cells on the accuracy of embryo classification (Figure 4); the panel for 5-cells in figure 4 is identical to Figure 3.

**Figure 4:**
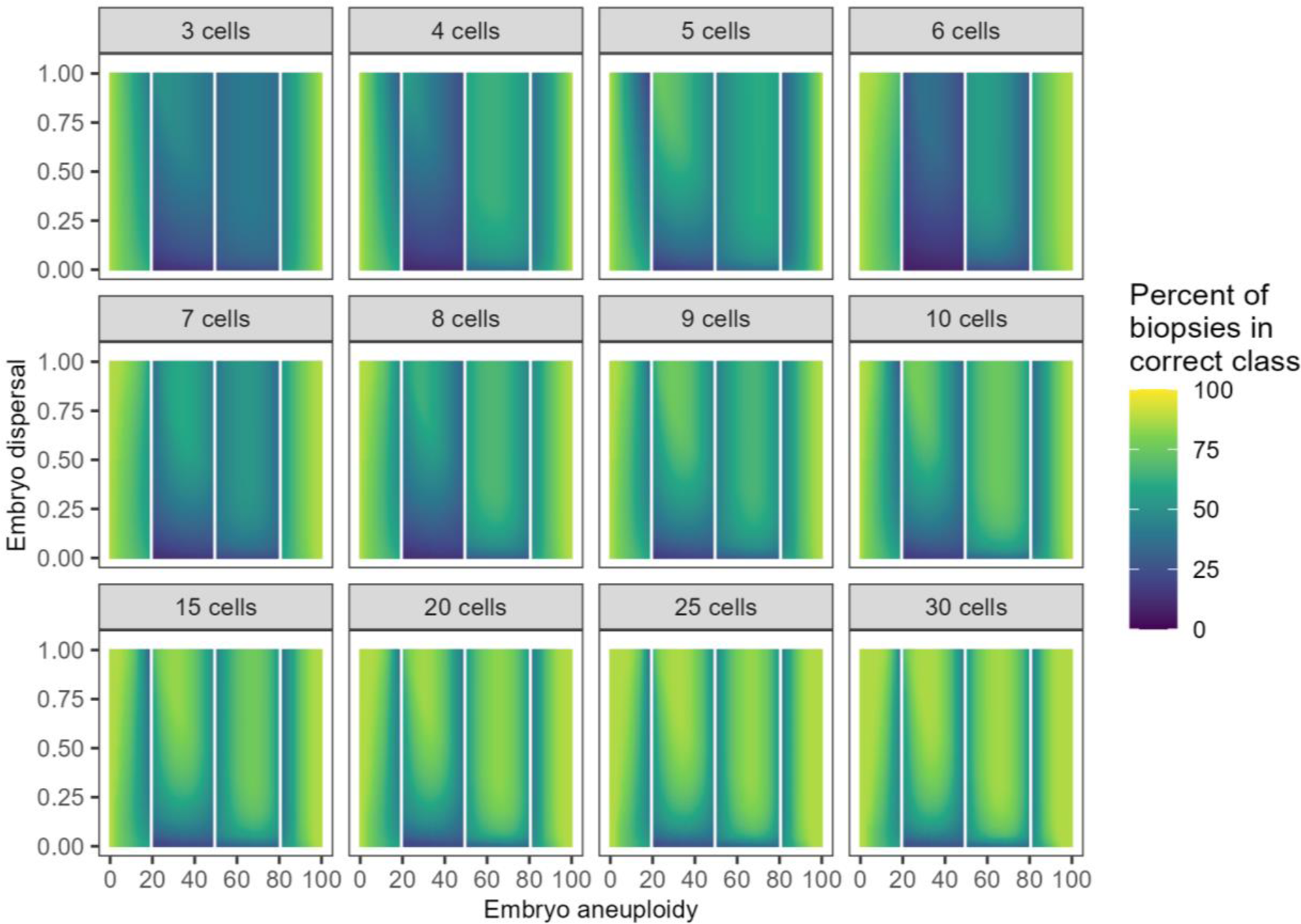
The effect of increasing biopsy size on accuracy for all combinations of embryo aneuploidy and dispersal. Numbers above each heatmap show the biopsy size. There are only small increases to accuracy over the 5-10 cell biopsy range, and even at larger biopsy sizes there is little difference to accuracy when dispersal of aneuploid cells is very low.

Notably, while accuracy does increase as biopsy size increases, we see little difference over a ‘practical’ biopsy size of 5-10 cells. We also see that even if the biopsy is unrealistically large (>15 cells), there is little improvement to biopsy accuracy in embryos with highly clustered aneuploid cells, especially in the low-level mosaic class. This suggests there is no clear benefit to biopsying larger numbers of cells. Indeed, the effect of using hard cut-off thresholds with these different biopsy sizes makes, for instance, a 6-cell biopsy appear less accurate over the high-level mosaic embryos than a 5-cell biopsy. Furthermore, classification accuracy of low- and high-level mosaic embryos is very poor for 3-cell biopsies and for 4-cell biopsies of low-level mosaic embryos, and we therefore caution against collecting fewer than 4 cells in a biopsy.

We then tested whether taking two 5-cell biopsies from an embryo improved over a single 10-cell biopsy. There is a small improvement for embryos with low dispersal (Figure S1-S3), increasing accuracy by up to ∼12 percentage points, but there are no benefits at higher dispersals.

### Predicting an embryo as high- or low-level mosaic from a single biopsy is not reliable

Up to this point, we have demonstrated how patterns of embryo biopsies vary as the embryo parameters change. We next considered the reverse situation, which is what clinicians actually experience: given a single biopsy, which embryos are most likely to generate this biopsy, and does this allow effective prediction of embryo status? For this, we made the assumption that an embryo is equally likely to originate from anywhere in the parameter space of aneuploidy and dispersal. This is probably not true of real embryos, but we do not have enough biological data to clearly constrain these values as yet (see the Discussion).

Under these assumptions, we simulated a pool of 1,020,100 embryos covering all pairwise combinations of aneuploidy and dispersal with 100 replicates per combination, and took all 202,020,000 possible biopsies from these embryos. We then grouped the biopsies by the number of aneuploid cells they contained. For each number of aneuploid cells, we counted how frequently biopsies with that number occurred in each embryo combination (Figure 5).

**Figure 5:**
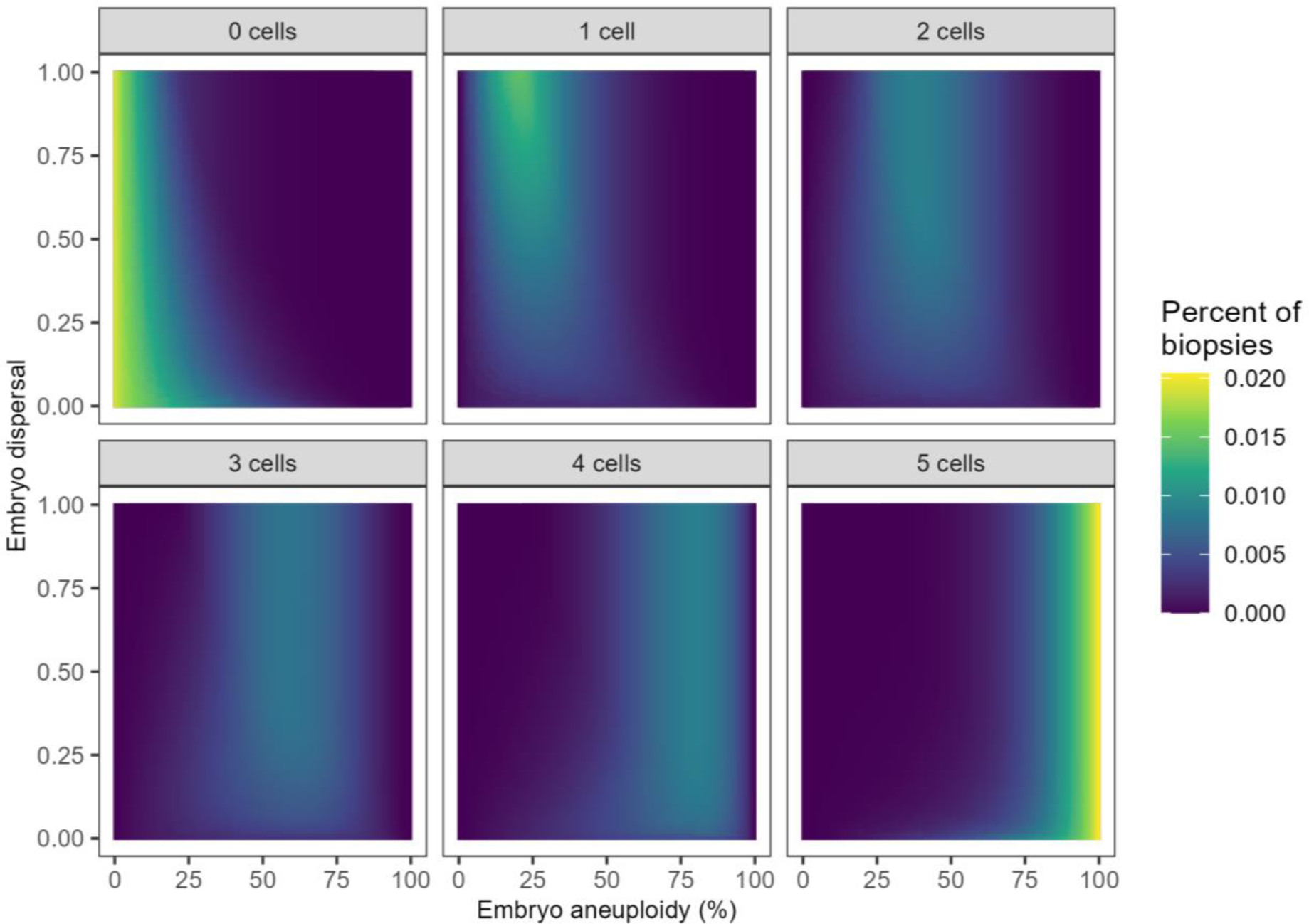
The possible origin embryos for 5-cell biopsies with 0 to 5 aneuploid cells. Biopsies with either 0 or 5 aneuploid cells are more likely to come from a constitutively normal or constitutively aneuploid embryo respectively, but mosaic biopsies can come from a much broader possible range of embryos.

We observed that while fully euploid or aneuploid biopsies are more likely to originate from correspondingly high or low aneuploid embryos, when biopsies are mosaic there is a wide range of possible origins. This shows that accurately classifying a mosaic embryo based on a single biopsy is not robust.

To demonstrate this further, we focussed on just three levels of dispersal - 0 (clustered), 0.5 (mid dispersal) and 1 (highly dispersed). For each level of biopsy aneuploidy, what percentage of biopsies originate at each level of embryo aneuploidy (Figure 6A)? In the ideal scenario biopsy aneuploidy and embryo aneuploidy would be perfectly correlated and we would be able to derive the precise level of aneuploid cells in a mosaic embryo for a given biopsy result. However, while there is a correlation, it is weak, especially at low dispersal (i.e. when cells are clustered). Adding the embryo classification thresholds on top of this chart, we then counted the number of embryos that matched the biopsy’s class (Figure 6B). At low dispersal, only about half of biopsies would correctly predict their embryo’s class, and even at high dispersal the accuracy rises to ∼80% only for fully euploid or aneuploid biopsies. A practical interpretation is that if one took a 5-cell biopsy from an embryo with clustered aneuploid cells, and the biopsy showed one aneuploid cell (low level mosaic), the embryo itself would be a low-level mosaic for less than half of such biopsies; even if the aneuploid cells were dispersed, the embryo would still be low-level mosaic for 55% of these biopsies. For a mosaic embryo, the classification would be as likely to be incorrect as correct.

**Figure 6:**
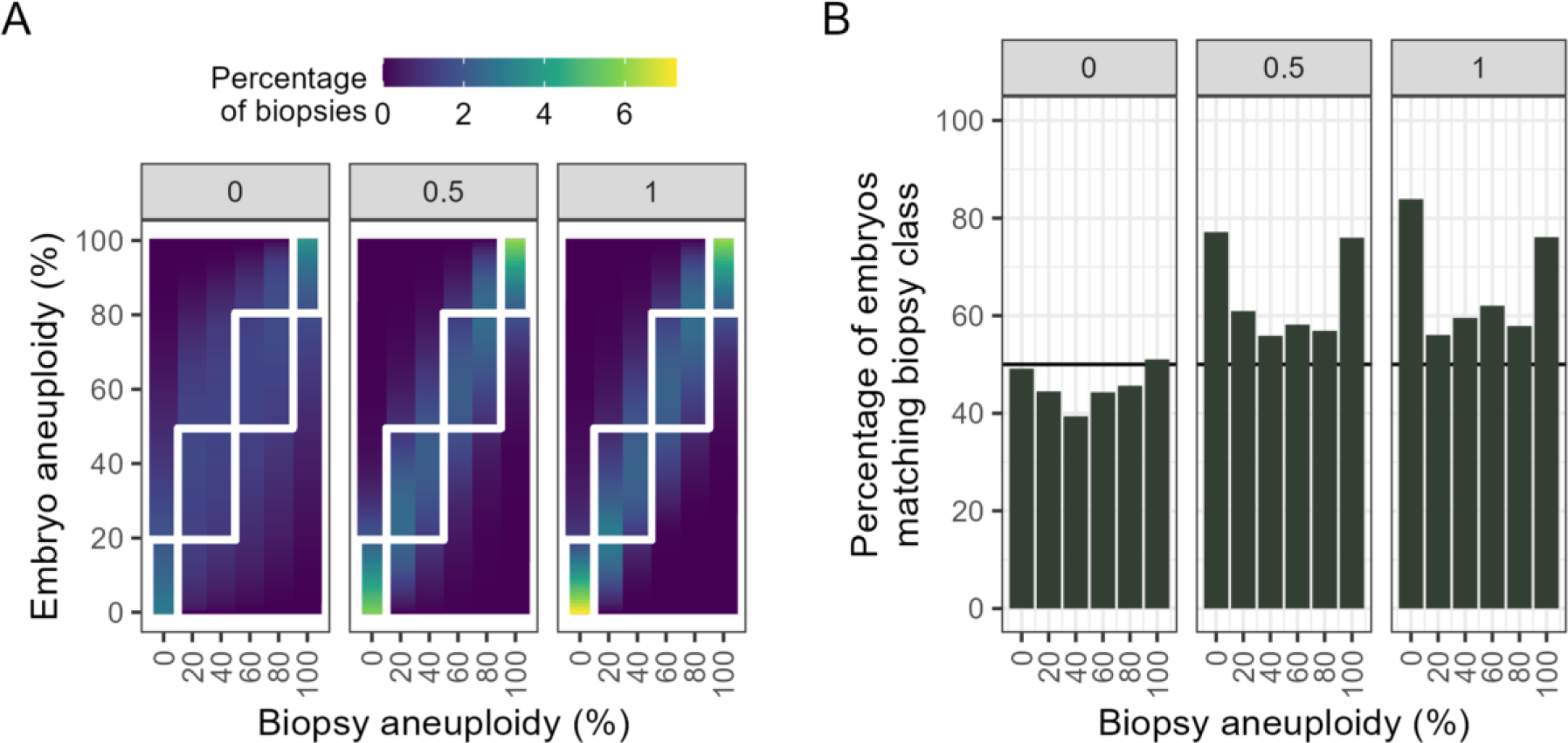
A mosaic biopsy has limited predictive power at classifying an individual embryo. (A) The percentage of biopsies with a given aneuploidy originating from an embryo with a given aneuploidy at three levels of dispersal (0, 0.5, 1). The embryo classification thresholds are drawn as white squares; for prediction to be useful, the majority of embryo aneuploidies should be within the squares. (B) the percentage of biopsies that correctly predict their embryo class at the three levels of dispersal.

### Biopsy results are nonetheless informative when ranking mosaic embryos

In clinical practice, it is rare to only consider one single biopsied embryo at a time. Usually multiple oocytes are retrieved following ovarian stimulation, of which some will be graded good quality and suitable for TE biopsy - *e.g.* 16.4±11 oocytes from which 4.9±4.7 blastocysts were biopsied and a single embryo transferred; (Lin et al., 2020). The clinical real-world question is “*which of these embryos (if any) should be transferred to yield the best chance for pregnancy?”*. The embryos are ranked, using the biopsies to determine their relative quality. Do the limitations imposed by single biopsy accuracy impair the ability to correctly rank two or more embryos?

Consider two embryos. One has 40% aneuploidy, and a biopsy yielding 2/5 aneuploid cells. The second has 60% aneuploidy and a biopsy yielding 3/5 aneuploid cells. The biopsies are *accurate* representations of the embryo. The biopsies can also be used to *rank* the embryos from less aneuploid to more aneuploid. Now consider the same embryos, but with different biopsy results: 0/5 and 2/5 aneuploid cells respectively. The biopsies are no longer *accurate*; they do not reflect the true level of aneuploidy in the embryo. However, they still correctly *rank* the embryos from less aneuploid to more aneuploid. Selecting the embryo with the lowest number of aneuploid cells in the biopsy for transfer is still the most sensible decision.

To understand how this ability to rank embryos is affected by aneuploidy and dispersal, we repeatedly generated two embryos with different aneuploidies, from 0-100%, took all possible biopsies from each embryo, and made all pairwise combinations of those biopsies. We scored the ranking as correct if the biopsy from the less aneuploid embryo had fewer aneuploid cells than the biopsy from the more aneuploid embryo. Equal numbers of aneuploid cells in the biopsies were scored as ties, and higher numbers of aneuploid cells in the biopsy from the less aneuploid embryo were scored as incorrect ranking.

We aggregated the rank results by the true difference in aneuploidies between the two embryos, since ranking embryos with a large aneuploidy difference should be more likely to succeed than ranking embryos with a small aneuploidy difference. Figure 7A shows the mean and standard deviation over 100 replicates for each aneuploidy combination at zero dispersal (the worst-case scenario for accuracy from our earlier modelling). Although some biopsies are tied (equal numbers of aneuploid cells), on average ranking embryos is successful more than half of the time as long as there is a greater than 25 percentage point difference in aneuploidies.

**Figure 7:**
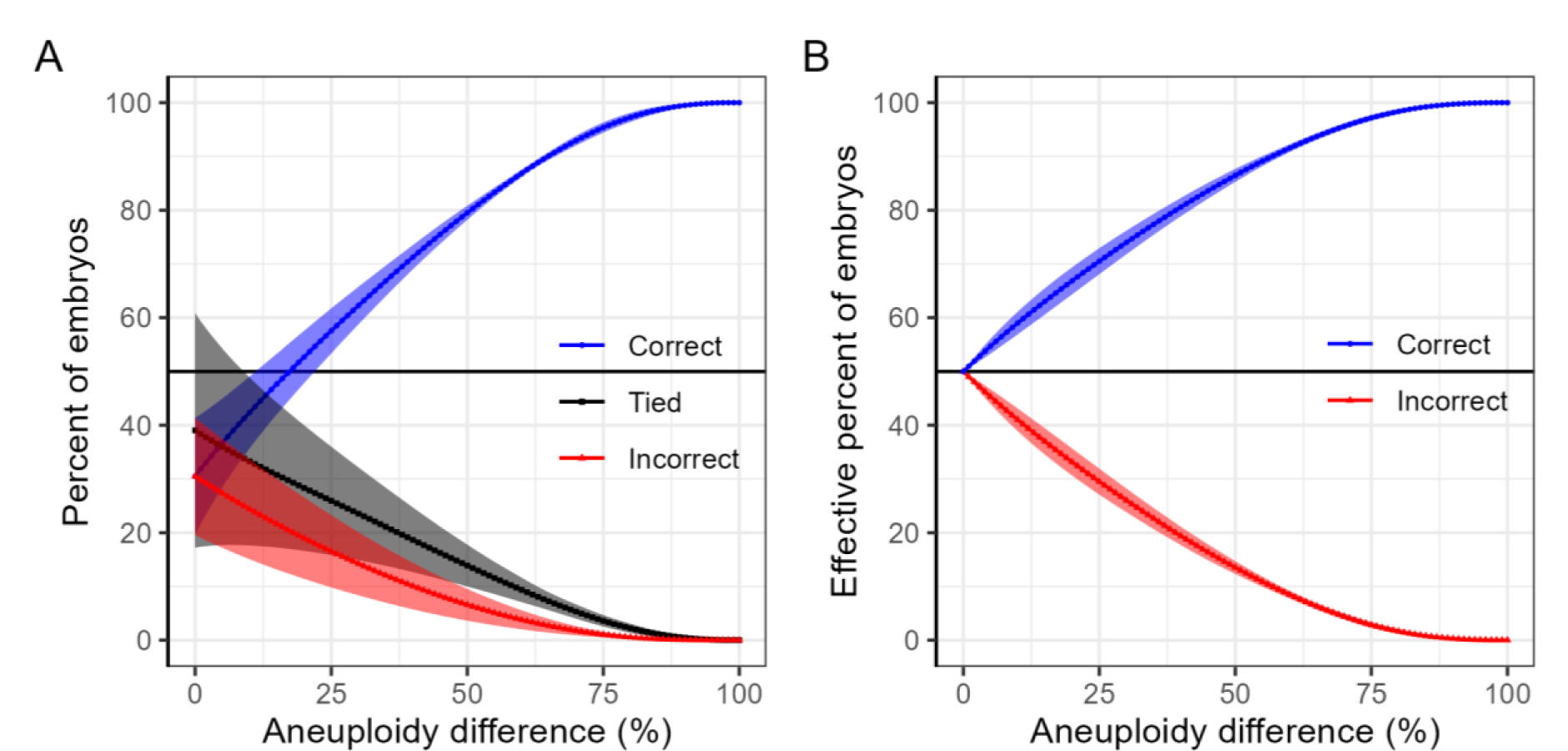
Effect of the size of aneuploidy differences on ranking two embryos **with different levels of aneuploidies** by biopsy result for embryos with zero dispersal. A) There is a greater than 50% chance of unambiguously choosing the correct rank order as long as the absolute difference in aneuploidy is greater than 25%. B) Biopsies with equal aneuploidy (tied ranking) are evenly split between correct and incorrect ranking to mimic the clinical scenario in which a random choice will be correct 50% of the time. Ranking two embryos based on biopsy outcomes will **always** on average correctly rank embryos better than chance if the embryos have unequal levels of aneuploidy. 100 replicate embryos were generated per aneuploidy combination. Values show mean and standard deviation after aggregating by aneuploidy difference. More detailed breakdowns by aneuploidy and dispersal combinations are shown in supplementary data (Figures S4 - S5).

What practically happens in the clinic if two embryos have equal biopsy results, and one is desired for transfer? The data must be either discarded as uninformative, and other embryo characteristics (*e.g.* morphology) relied upon, or a random choice must be made between the two embryos. In our simulated embryo pairs, a random choice would correctly select the embryo with lower aneuploidy half the time. We accounted for this by splitting half of the ‘Tied’ rank biopsies each into the ‘Correct’ and ‘Incorrect’ ranks (Figure 7B), yielding the effective ranking ability. As long as there is a difference in aneuploidy between two embryos, no matter how small, biopsy data will on average allow the two to be distinguished better than by chance. This demonstrates that biopsying an embryo is useful for comparisons *between* embryos, *even if it is not directly informative as to the absolute aneuploidy status of the embryo*.

In practice, clinics are not only trying to distinguish between two embryos alone. The goal is often to select the best **k** embryos from a pool of **n**. We simulated an embryo pool with random aneuploidy levels, from which three embryos were selected via single biopsy (Figure 8) and compared the selected embryos to the true “best” embryos. This ranking is effective across the full range of possible dispersals, and at other pool sizes and selection sizes (Figure S6).

**Figure 8:**
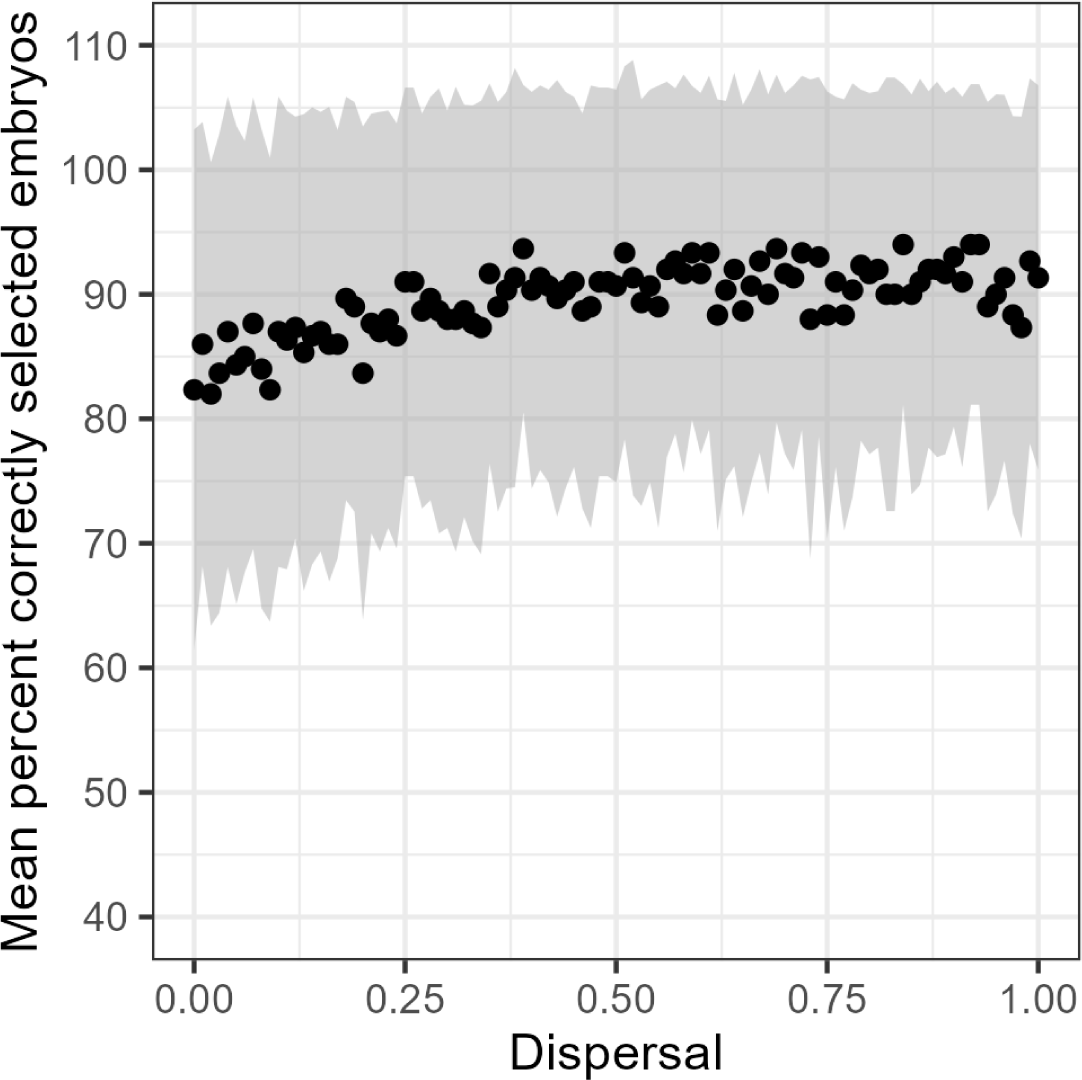
Single biopsies can reliably select the best three embryos from a pool of six embryos. For each dispersal level, 100 pools of embryos were generated with random aneuploidy levels. A single biopsy from each embryo was used to rank the pool, and the three embryos with lowest rank were selected. The figure shows the mean percentage of selected embryos that are in the true “best three”. Values show mean and standard deviation from 100 embryo pools. Full combinations are in Figure S6.

### Embryo transfer data demonstrate association of aneuploidy level with clinical outcomes

Viotti et al. (2021) provided an analysis of outcome data from 1000 mosaic embryos showing statistically significant inverse correlation between aneuploidy level and favourable outcomes (implantation as measured via gestational sac, ongoing pregnancy and birth). Here, we extend that data with an additional 733 mosaic embryos. In confirmation of the previous analysis, we show there is still a significant difference in both implantation and ongoing pregnancy / birth outcomes between mosaic embryos classified as less than 50% or greater than 50% aneuploid (Figure 9).

**Figure 9:**
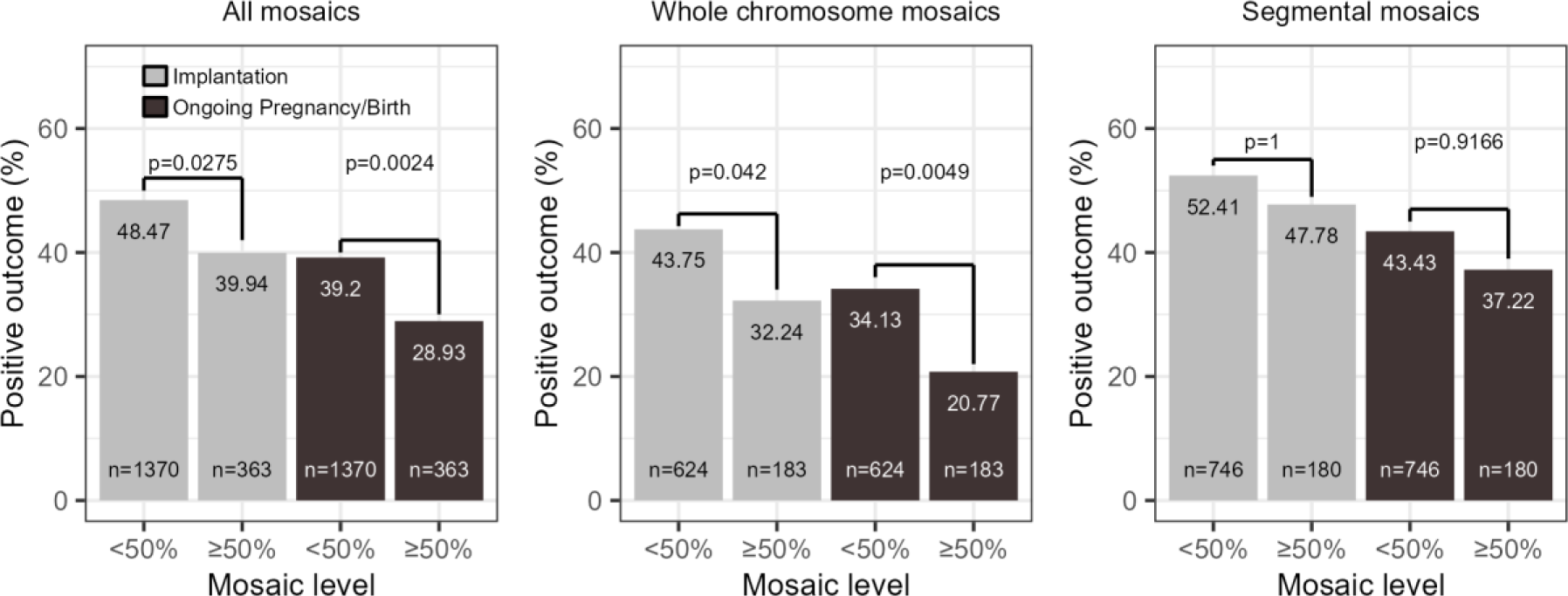
Implantation (light grey) and ongoing pregnancy/birth rates (dark grey) split by segmental or whole chromosome aneuploidies. Complex mosaics with multiple changes detected were classified based on the largest abnormality present. Thus ‘Whole chromosome mosaics’ includes all cases involving a full chromosome aneuploidy regardless of the presence or absence of additional segmental aneuploidies, while ‘Segmental mosaics’ includes only cases where all aneuploid regions were sub-chromosomal in extent. Breaking the data into <50% or ≥50% aneuploidy reveals significant differences in outcome for whole chromosome mosaics, though not for segmental mosaics (chi square tests with Bonferroni correction). This figure is structured to match Viotti et al. 2021 Figure 2A for comparison.

We used logistic regression to further analyse the relationship between aneuploidy level and aneuploidy type on the probability of successful outcome of implantation or ongoing pregnancy/birth (Figure S8). We found that, holding aneuploidy type constant, the odds of successful implantation decreased by 1.14% (Odds Ratio [OR]: 0.989, CI: 0.981-0.996) for each additional percentage of aneuploidy. Holding aneuploidy level constant, the odds of successful implantation decreased by 33.9% (OR: 0.661, CI: 0.554 - 0.799) for mosaics with whole chromosomal aneuploidies compared to segmental aneuploidies.

For ongoing pregnancy / births, holding aneuploidy type constant, the odds of successful outcome decreased by 1.50% (OR: 0.985, CI: 0.977-0.993) for each additional percentage of aneuploidy. Holding aneuploidy level constant, the odds of successful outcome decreased by 38.2% (OR: 0.618, CI: 0.506 - 0.754) for mosaics with whole chromosomal aneuploidies compared to segmental aneuploidies.

## Discussion

The chromosomally complex, fluid and dynamic nature of the human embryo (Coticchio et al., 2021), coupled with the widespread use of PGT-A make it no surprise that it is still an area of great debate. The data presented here provide evidence to resolve an apparent paradox on whether the practice of PGT-A is effective when embryos are mosaic. Our modelling demonstrates that mosaic biopsies can derive from a wide range of embryos and their results should not be considered a reliable indicator of individual embryo status. Somewhat counterintuitively however, when considering a pool of embryos that need to be ranked based on the proportion of aneuploid (vs euploid) cells, a mosaic biopsy result can be informative in establishing that ranking. Furthermore, when a biopsy indicates that all cells are aneuploid, or all cells are euploid, it is considerably more informative in predicting the genetic status of the embryo.

### Our modelling supports PGT-A outcome data

The data presented here support the outcome data in the mosaicism registry (1733 embryos at the time of writing) suggesting that the proportion of aneuploid cells in a mosaic biopsy is a predictor of the chances of live birth – the greater the level of aneuploidy in the mosaic biopsy, the lower the chances of a live birth. This also supports data from post-2020 non-selection trials and the unblinded cohort study of Gleicher and colleagues (Barad et al., 2022; Tiegs et al., 2021; Wang et al., 2021; Yang et al., 2021), in which embryo biopsies diagnosed as 100% euploid have a higher chance of pregnancy and live birth (55-65%) than any classes in the mosaicism registry. On the other hand, those diagnosed as 100% aneuploid have little or no chance (0-2%). Of the 4 studies that have addressed this issue, a total of 267 embryos have been transferred, with only 3 (1%) leading to chromosomally normal live births (in 2 of the 4 studies the live birth rate was 0%). The three surviving births presumably represented genotyping errors, or mosaic embryos in which a postzygotic error led to a cluster of aneuploid cells around the site of biopsy.

The debate over the utility of PGT-A often centres around the compounding effects of mosaicism and the interpretation of its results. While there are few randomised controlled trials (RCTs) for PGT-A, meta-analysis of RCTs has suggested chromosome copy number screening increases implantation rates (Dahdouh et al., 2015). More recent RCTs suggest that PGT-A does not increase cumulative live birth rate in women between 20 and 37 (average age 29) (Yan et al., 2021) but these studies may be under-powered (Cornelisse et al., 2020). Data is especially lacking on the utility of PGT-A for women over 35, a high-risk group for aneuploidies. Munné et al., (2019) found no increase in ongoing pregnancy rate (OPR) at 20 weeks in women aged 25-40 associated with PGT-A, though post-hoc analysis of women aged 35-40 showed a significant increase in OPR. In light of the lack of conclusive RCTs and biological uncertainty about the impacts of mosaicism, Gleicher et al., (2021) have suggested PGT-A should not be routinely offered in the clinic, as it risks discarding viable embryos, does not provide reliable information and may give clinics a conflict of interest in recommending expensive yet unnecessary treatments to patients (Gleicher et al., 2021, 2020).

Most data for higher risk groups come from published pregnancy rates from clinics and deal with pregnancy or live birth rates per embryo transfer. It is very clear that, by this measure, PGT-A confers a benefit (Sanders et al., 2021), particularly in older women where the difference is over tenfold. Such results are borne out by other non-selection trials (Tiegs et al., 2021), and an unblinded cohort study (Barad et al., 2022) in which fully aneuploid diagnoses were transferred, but only led to live birth 1% of the time. Strong support for the effectiveness of biopsies in selecting mosaic embryos for transfer has come from analysis of outcomes (Viotti et al., 2021) and our follow-up analysis here, which show clear differences in rates of implantation and ongoing pregnancy for embryos with low level or high-level mosaicism. Given the good/poor prognosis for constitutively euploid/aneuploid embryos respectively, this is of primary importance for cycles where *only* embryos with a mosaic diagnosis. The proportion of such cycles is not well characterised but has been reported to be up to 20% (Lin et al., 2020).

### Relevance for clinical interpretation of biopsy data

The modelling presented here provides evidence that biopsy results (even mosaic ones) can be interpreted with some confidence when ranking embryos for potential transfer, but give limited information about a single embryo. Furthermore, assigning a classification to a biopsy such as “euploid, low-level mosaic, high-level mosaic or aneuploid” can also affects the accuracy of the reporting. In the light of the results presented here we respectfully suggest that while “euploid and aneuploid” should be retained for biopsies showing 0% or 100% aneuploid cells respectively, the classification of mosaic embryos based on biopsies with intermediate levels of aneuploidy detection is less robust. Clinicians should bear in mind that simple fixed cut-offs for “high” or “low” levels of mosaicism can be misleading when considering only a single embryo. Rather, we suggest using classification or absolute aneuploidy level preferentially for ranking embryos, regardless of any nominal cutoffs. We have shown that any absolute difference in the measured aneuploidy level is sufficient to rank one embryo as higher or lower grade aneuploidy than another at better than chance accuracy, though if the measured aneuploidy levels for two embryos fall within 20% of each other, clinicians may wish to weight other aspects of embryo quality (such as morphology) more highly when selecting embryos for transfer. Our testing of biopsy sizes showed classification accuracy of low- and high-level mosaic embryos is poor for 3-cell biopsies and for 4-cell biopsies of low-level mosaic embryos and we therefore suggest collecting at least 5 cells if possible. We also found only a small benefit to taking two 5-cell biopsies versus one 10-cell biopsy, and thus given the robust ranking of embryos in a pool from single biopsies, we do not recommend multiple biopsies.

### Future extensions to modelling for consideration

In this study, while we addressed the level of aneuploidy in the biopsy and its relationship to the likely level of aneuploidy in the whole embryo, we did not consider that the TE forms the placenta and other extraembryonic tissue, distinct from the inner cell mass (ICM) that will develop into the foetus. However, TE biopsy reflects the chromosomal constitution of the ICM in studies from both humans (Ren et al., 2022; Victor et al., 2019) and cattle (Tutt et al., 2021), and uniform aneuploidies are well detected by TE biopsy (Popovic et al., 2019; Victor et al., 2019). We also do not consider ‘self-correction’; the long-established phenomenon that cleavage stage embryos (day 3) can have aneuploidy in more than half of their cells but that level nonetheless declines by trophectoderm stage (day 5) (reviewed in McCoy, 2017).

Of all the parameters explored (aneuploidy, dispersal, embryo size, biopsy size), we found that the level of dispersal is the most important factor affecting the accuracy of a biopsy. If the aneuploid cells are evenly dispersed across the embryo, a biopsy may be fully representative, but if the aneuploid cells are clustered, individual biopsies are likely to be misleading. The level of dispersal may also be influenced by the type of error generating the mosaicism in the first place; a nondisjunction error at meiosis followed by a trisomy rescue event may have a different pattern of aneuploidies to a polymerase slippage at mitosis, or a mitotic nondisjunction.

While our model has considered many parameter combinations, it has also still been using a binary treatment of aneuploidy: a cell is aneuploid or it is euploid. In reality, aneuploidies can be more complex, with different chromosomes having different likelihoods of aneuploidy, and an embryo may contain multiple different karyotypes. Aneuploidies can also be segmental, affecting regions of chromosomes and not entire chromosomes. We see from the clinical outcome data analysed here that segmental mosaics do not have as strong a negative impact on outcomes as whole chromosome mosaics. Aneuploidies may also be balanced, invisible to bulk sequencing, and only revealed by single cell sequencing (Ren et al., 2022), and mitotic nondisjunction events typically generate trisomic and monosomic cells that could balance each other out and thus not easily be detectable by NGS when pools of cells are analysed. We want to investigate these additional aspects of mosaicism in the future and how they can affect the measured biopsy outcomes. The true incidence of different levels of embryo aneuploidy and dispersal - both in natural populations and in the patient groups that are typically undergoing PGT-A is also not fully understood. Only with real data on the distribution of aneuploidies and their dispersal in embryos will we be able to refine these models. This can be obtained through (*e.g.*) lineage tracing experiments or more detailed sampling and single cell sequencing of embryos that have been rejected for transfer.

### Conclusion

The assisted reproduction community has been debating the merits of PGT-A since its inception and, in our opinion, our data provide a resolution to some of the opposing viewpoints around mosaicism. While debate on other aspects of PGT-A will continue, we suggest a combination of the mosaicism registry data with the modelling presented in this study should inform the decision-making process when a mosaic result is returned from an embryo biopsy. What we have demonstrated is that although a biopsy result is imperfect, it is nonetheless informative in prioritising embryos for transfer whether a 100% euploid, 100% aneuploid or mosaic result is obtained. When navigating hazards, even a blurred map is useful.

## Data Availability

Scripts used perform the modelling in this paper are available at https://github.com/bmskinner/embryo_biopsy. IRMET data is available upon request.

## Acknowledgements

The authors acknowledge the use of the High-Performance Computing Facility (Ceres) and its associated support services at the University of Essex in the completion of this work.

## Funding

BMS is supported by HEFCE/UKRI funding (University of Essex). PJIE is supported by HEFCE/UKRI funding (University of Kent).

## Competing interests

DKG consults for, and has ongoing projects with, Care Fertility, Igenomix, Conceivable, Cooper and London Women’s Clinic. BMS, MV and PJIE have no competing interests to declare.

## Data Availability

Scripts used to generate analyses in this paper are available at https://github.com/bmskinner/embryo_biopsy. The tessera package is available under the open source GPL-3.0 license at https://github.com/bmskinner/tessera. IRMET data is available on request.

## Authors’ Contributions

Conceptualisation, BMS, PE and DKG; Methodology, BMS and PE; Software and Validation, BMS; Investigation, BMS and PE; Data Curation and Formal Analysis, BMS; Visualisation, BMS; Supervision and Project Administration, BMS and DKG; Writing - Original Draft, BMS, DKG and PE; Writing - Review and Editing, BMS, DKG, MV and PE; Resources, BMS and MV. All authors gave final approval for publication.

## Supplementary figures

**Figure S1:**
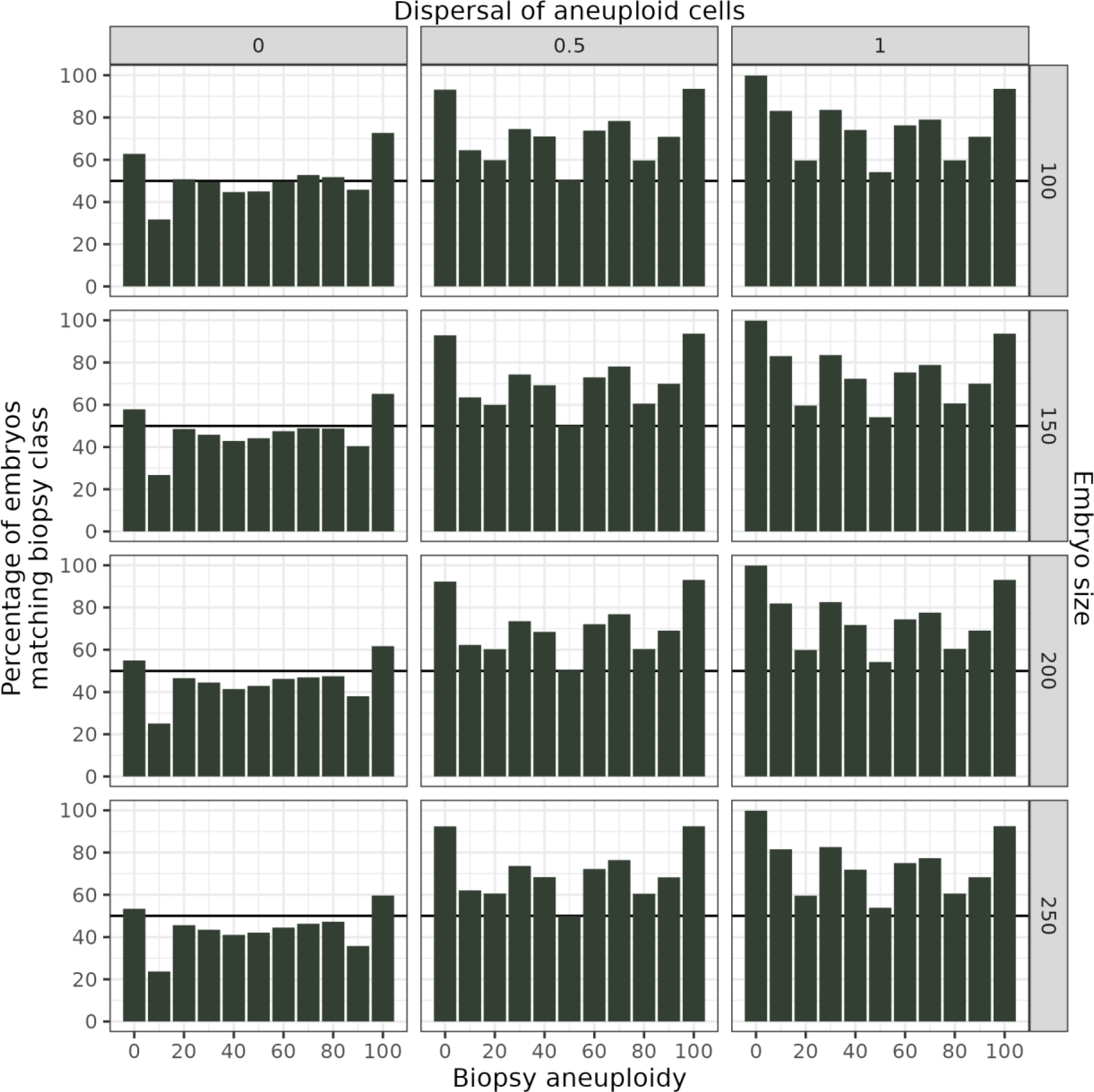
Accuracy of one 10-cell biopsy across embryo sizes and dispersals. Compare to Figure 6 (one 5-cell biopsy) and Figures S2 and S3 (two 5-cell biopsies).

**Figure S2:**
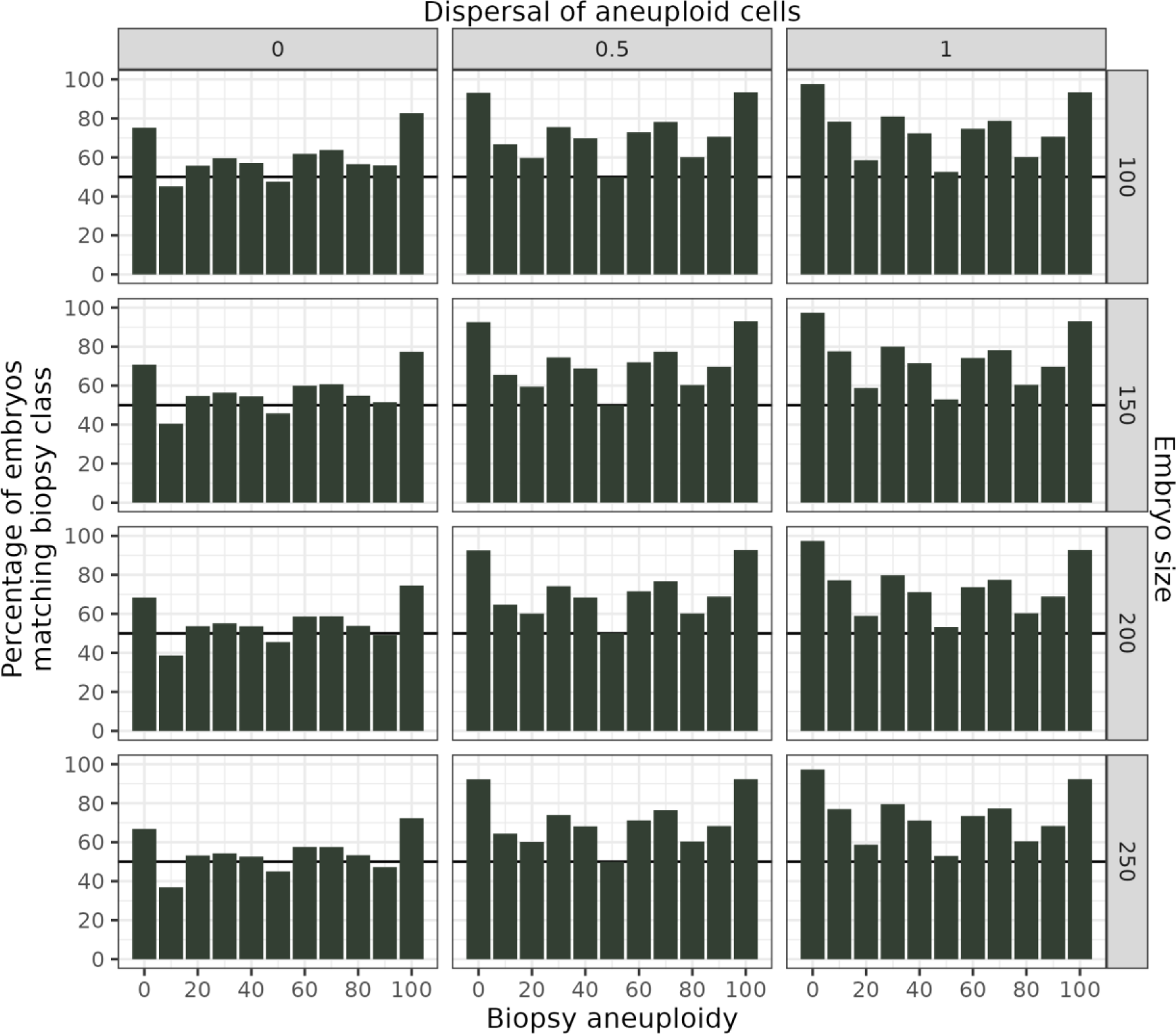
Percentage of two 5-cell biopsies matching their embryo class embryo sizes and dispersals. Compare to the single 5-cell biopsy in Figure 6B and the single 10-cell biopsy in Figure S1.

**Figure S3:**
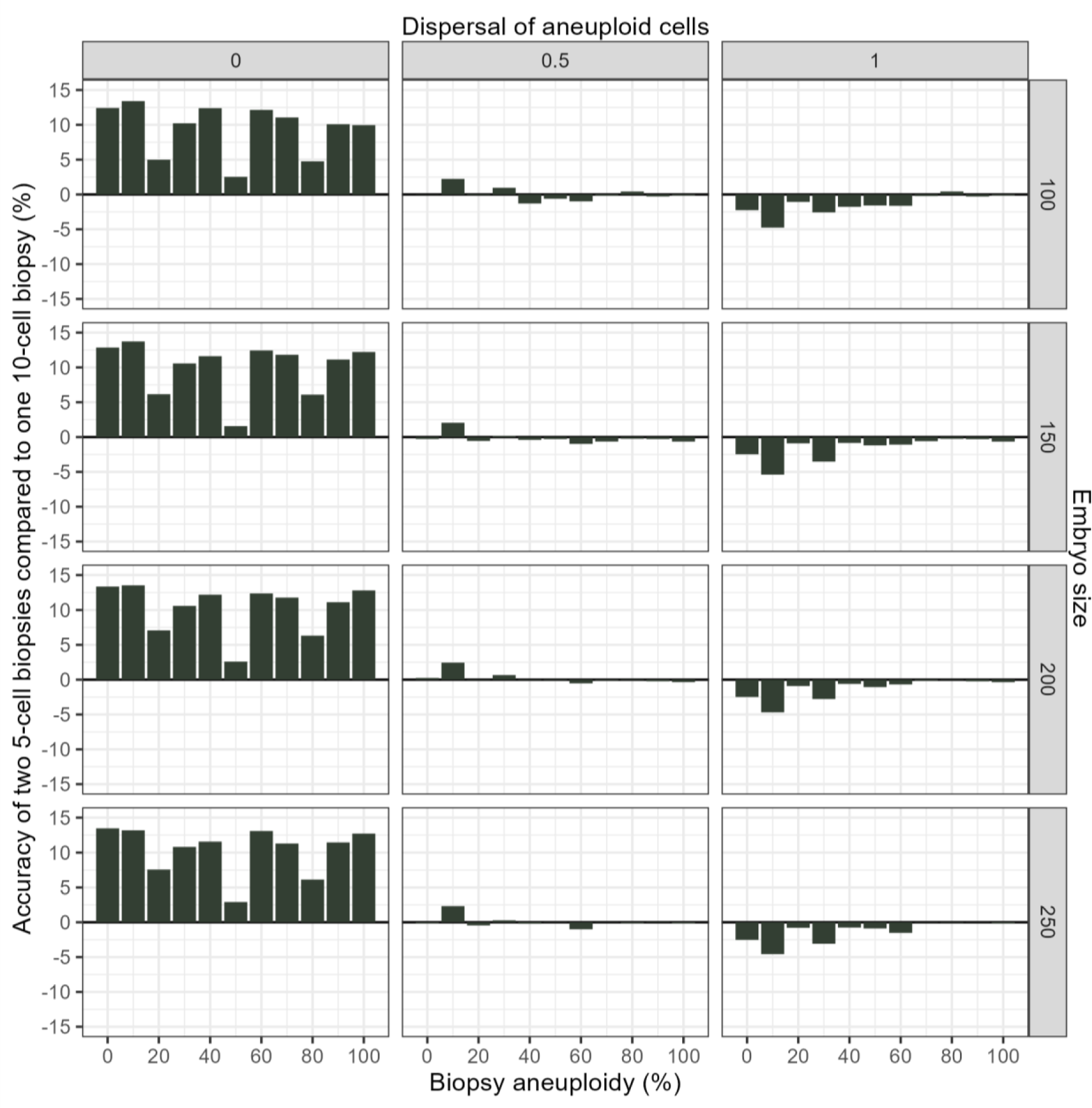
Comparison of one 10-cell biopsy and two 5-cell biopsies. Accuracy is increased with two 5-cell biopsies for embryos with low dispersal by up to ∼12 percentage points. There are no benefits at higher dispersals. This chart is the difference between Figures S1 and S2.

**Figure S4:**
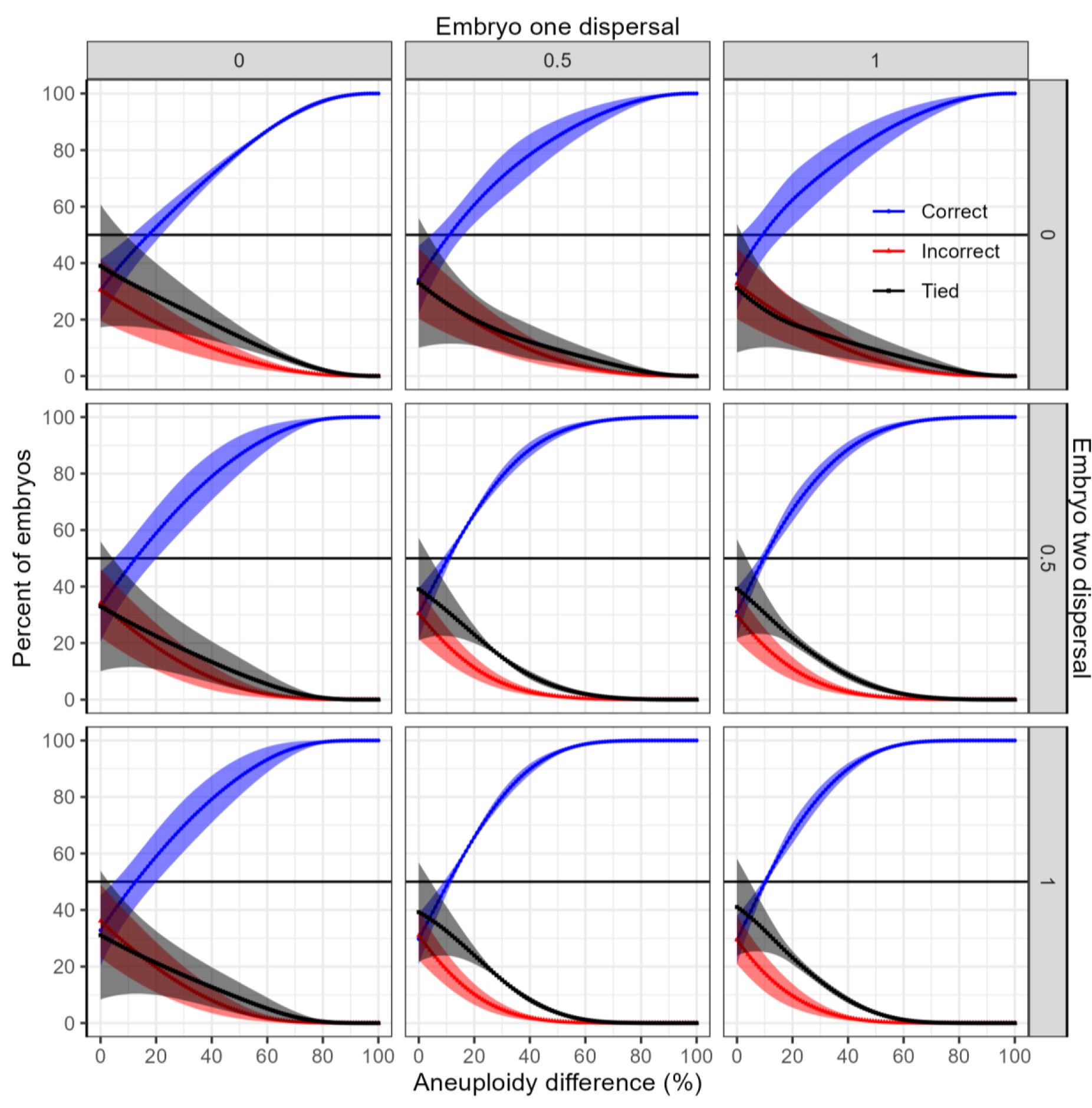
Effect of dispersal and aneuploidy differences on ranking two embryos with different aneuploidies by biopsy result. There is a greater than 50% chance of choosing the correct rank order at all levels of dispersion as long as the absolute difference in aneuploidy is greater than 20%. 100 replicate embryos were generated per aneuploidy and dispersal combination. Values show mean and standard deviation after aggregating by aneuploidy difference. Top left panel is equivalent to Fig 7A.

**Figure S5:**
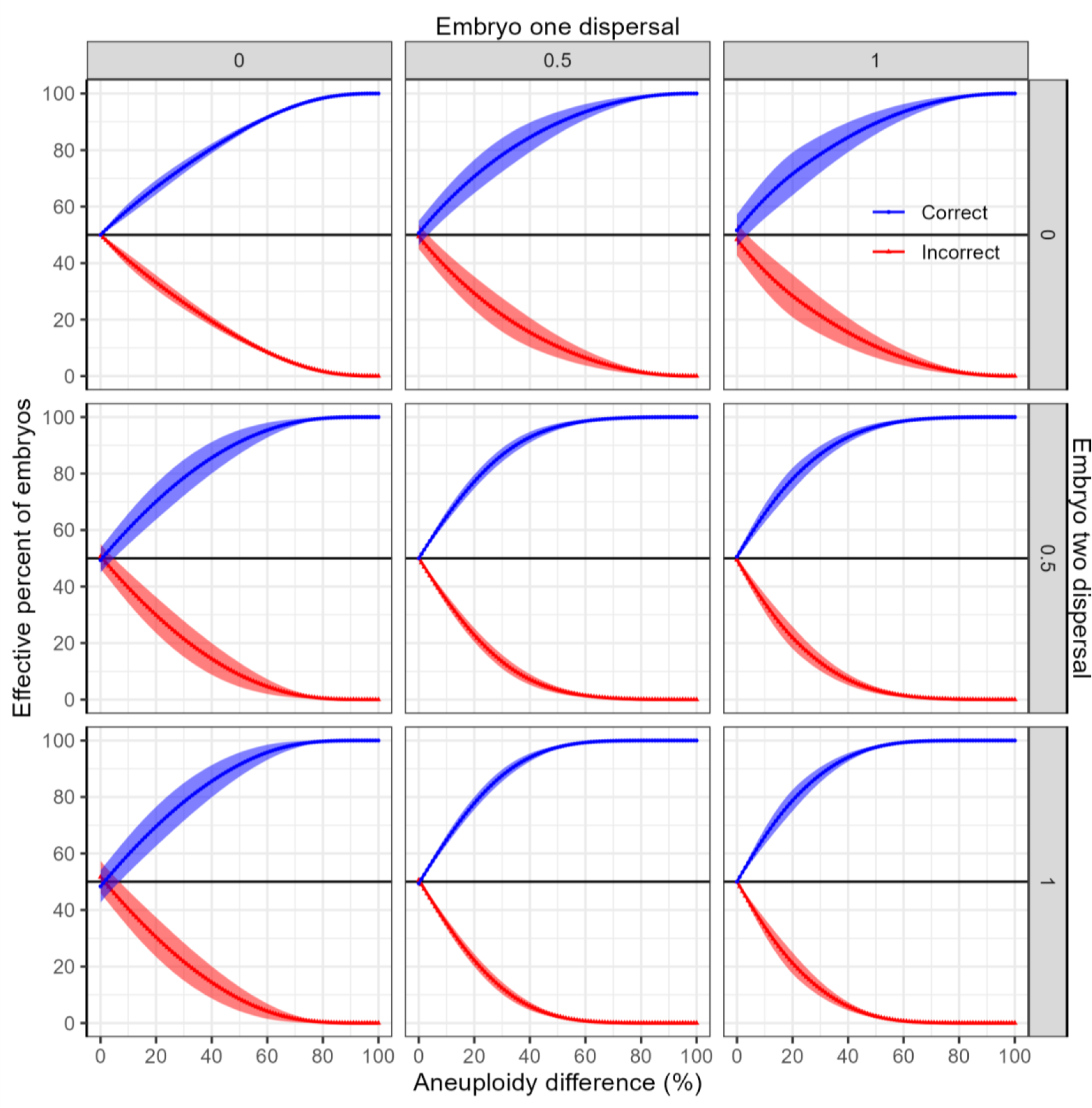
Contrast with Fig S4. Effect of dispersal and aneuploidy differences on ranking two embryos with different aneuploidies by biopsy result. Biopsies with equal aneuploidy (tied ranking) have been evenly split between correct and incorrect ranking, since a random choice will be correct 50% of the time. Ranking two embryos based on biopsy outcomes will always on average rank embryos better than chance if the embryos have unequal levels of aneuploidy. Top left panel is equivalent to Fig 7B.

**Figure S6:**
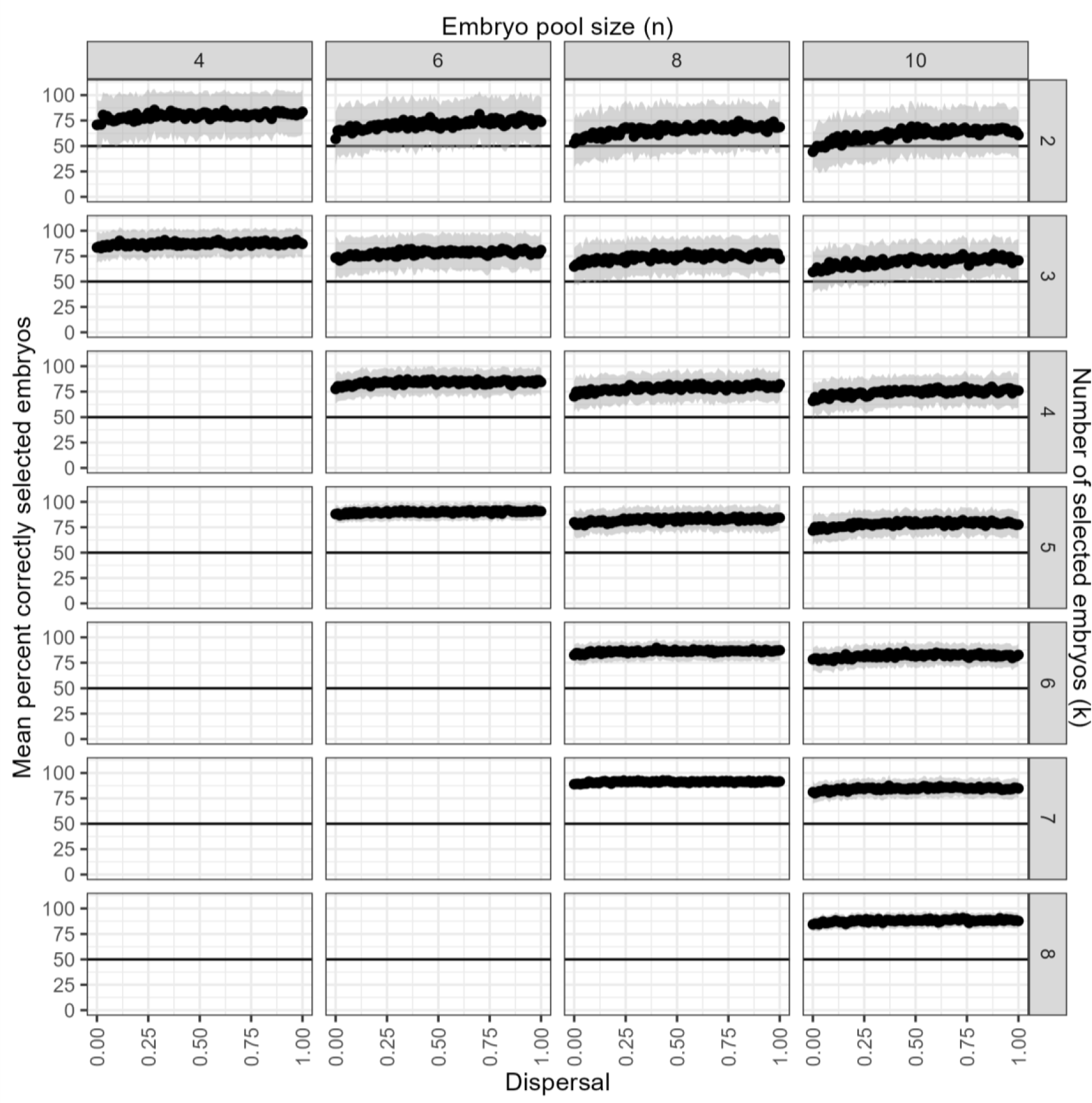
The ability to select the best k embryos from a pool of n when the pool contains embryos with random levels of aneuploidies. Embryos are correctly selected better than chance in almost all cases (the exception at large pool sizes and low selection sizes). All embryos in the pool had 200 cells. Values are mean and standard deviation of 100 replicates. This figure complements Figure 8.

**Figure S7:**
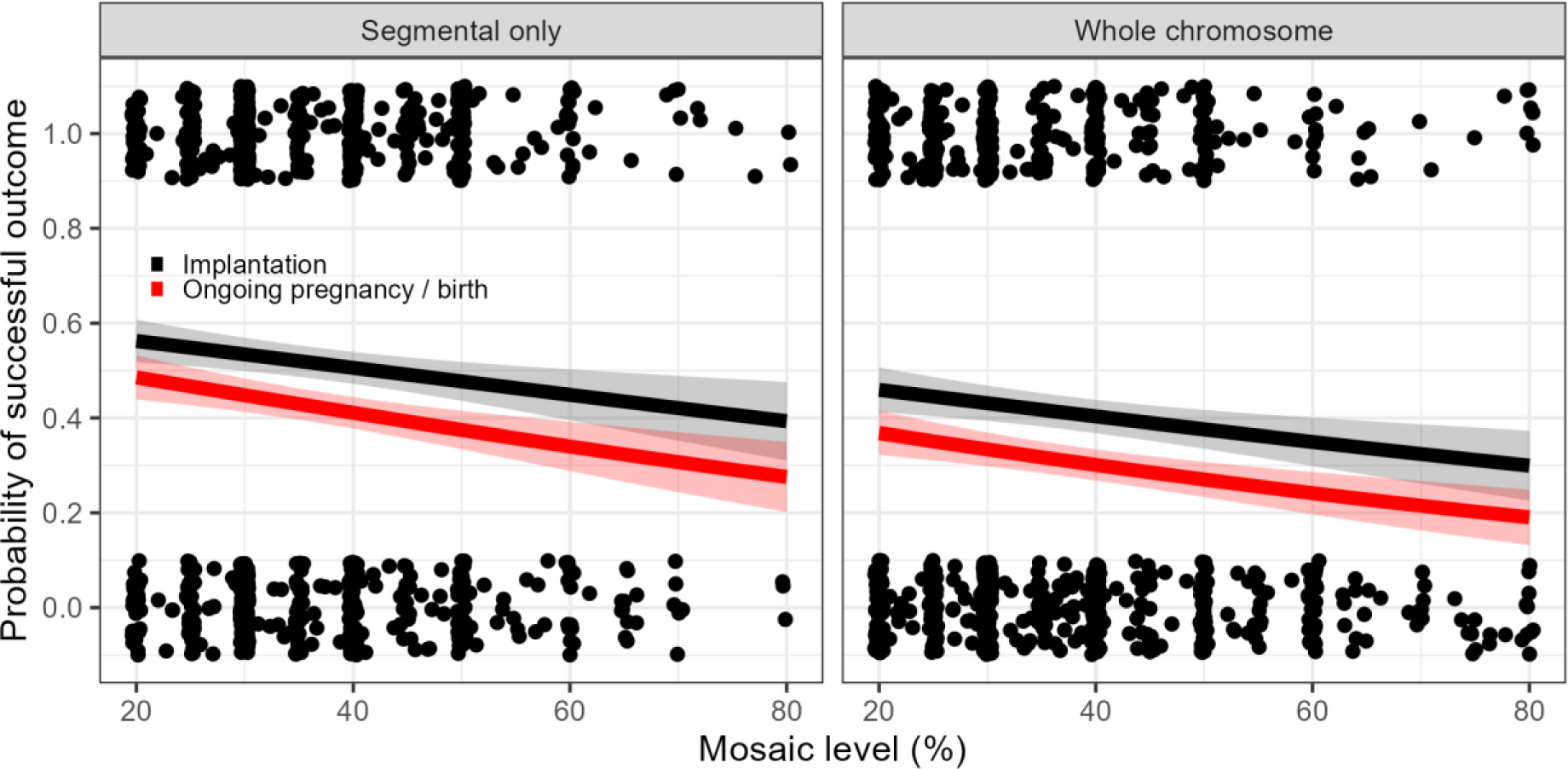
Logistic regression on outcomes shows a significant association with aneuploidy level and with the type of chromosomal abnormality. Points show the individual embryos, and are jittered for clarity. Lines show predicted outcomes for a given aneuploidy level from the logistic regression models and the shaded areas show the standard error. Both aneuploidy type and aneuploidy level have significant contributions to outcomes.

## Notes

### Author Declarations

Analyses of clinical data were approved by the IRB of the Zouves Foundation (OHRP IRB00011505, Protocol #0002).

## References

Barad DH, Albertini DF, Molinari E, Gleicher N. 2022. IVF outcomes of embryos with abnormal PGT-A biopsy previously refused transfer: a prospective cohort study. Human Reproduction 37:1194–1206. doi:10.1093/humrep/deac063

Chang W, Cheng J, Allaire JJ, Sievert C, Schloerke B, Xie Y, Allen J, McPherson J, Dipert A, Borges B. 2022. shiny: Web Application Framework for R.

Cornelisse S, Zagers M, Kostova E, Fleischer K, van Wely M, Mastenbroek S. 2020. Preimplantation genetic testing for aneuploidies (abnormal number of chromosomes) in in vitro fertilisation. Cochrane Database Syst Rev 9:CD005291. doi:10.1002/14651858.CD005291.pub3

Coticchio G, Barrie A, Lagalla C, Borini A, Fishel S, Griffin D, Campbell A. 2021. Plasticity of the human preimplantation embryo: developmental dogmas, variations on themes and self-correction. Human Reproduction Update 27:848–865. doi:10.1093/humupd/dmab016

Dahdouh EM, Balayla J, García-Velasco JA. 2015. Comprehensive chromosome screening improves embryo selection: a meta-analysis. Fertility and Sterility 104:1503–1512. doi:10.1016/j.fertnstert.2015.08.038

Gleicher N, Albertini DF, Barad DH, Homer H, Modi D, Murtinger M, Patrizio P, Orvieto R, Takahashi S, Weghofer A, Ziebe S, Noyes N. 2020. The 2019 PGDIS position statement on transfer of mosaic embryos within a context of new information on PGT-A. Reprod Biol Endocrinol 18:57. doi:10.1186/s12958-020-00616-w

Gleicher N, Metzger J, Croft G, Kushnir VA, Albertini DF, Barad DH. 2017. A single trophectoderm biopsy at blastocyst stage is mathematically unable to determine embryo ploidy accurately enough for clinical use. Reprod Biol Endocrinol 15:33. doi:10.1186/s12958-017-0251-8

Gleicher N, Patrizio P, Brivanlou A. 2021. Preimplantation Genetic Testing for Aneuploidy - a Castle Built on Sand. Trends Mol Med 27:731–742. doi:10.1016/j.molmed.2020.11.009

González Á. 2009. Measurement of Areas on a Sphere Using Fibonacci and Latitude– Longitude Lattices. Math Geosci 42:49. doi:10.1007/s11004-009-9257-x

Greco E, Minasi MG, Fiorentino F. 2015. Healthy Babies after Intrauterine Transfer of Mosaic Aneuploid Blastocysts. N Engl J Med 373:2089–2090. doi:10.1056/NEJMc1500421

Griffin DK, Brezina PR, Tobler K, Zhao Y, Silvestri G, Mccoy RC, Anchan R, Benner A, Cutting GR, Kearns WG. 2022. The human embryonic genome is karyotypically complex, with chromosomally abnormal cells preferentially located away from the developing fetus. Hum Reprod 38:180–188. doi:10.1093/humrep/deac238

Griffin DK, Ogur C. 2018. Chromosomal analysis in IVF: just how useful is it? Reproduction 156:F29–F50. doi:10.1530/REP-17-0683

Harton GL, Cinnioglu C, Fiorentino F. 2017. Current experience concerning mosaic embryos diagnosed during preimplantation genetic screening. Fertility and Sterility 107:1113– 1119. doi:10.1016/j.fertnstert.2017.03.016

Kim J, Tao X, Cheng M, Steward A, Guo V, Zhan Y, Scott RT, Jalas C. 2022. The concordance rates of an initial trophectoderm biopsy with the rest of the embryo using PGTseq, a targeted next-generation sequencing platform for preimplantation genetic testing-aneuploidy. Fertility and Sterility 117:315–323. doi:10.1016/j.fertnstert.2021.10.011

Leigh D, Cram DS, Rechitsky S, Handyside A, Wells D, Munne S, Kahraman S, Grifo J, Katz-Jaffe M, Rubio C, Viotti M, Forman E, Xu K, Gordon T, Madjunkova S, Qiao J, Chen Z-J, Harton G, Gianaroli L, Simon C, Scott R, Simpson JL, Kuliev A. 2022. PGDIS position statement on the transfer of mosaic embryos 2021. Reproductive BioMedicine Online. doi:10.1016/j.rbmo.2022.03.013

Lin P-Y, Lee C-I, Cheng E-H, Huang C-C, Lee T-H, Shih H-H, Pai Y-P, Chen Y-C, Lee M-S. 2020. Clinical Outcomes of Single Mosaic Embryo Transfer: High-Level or Low-Level Mosaic Embryo, Does It Matter? Journal of Clinical Medicine 9:1695. doi:10.3390/jcm9061695

Mantikou E, Wong KM, Repping S, Mastenbroek S. 2012. Molecular origin of mitotic aneuploidies in preimplantation embryos. *Biochimica et Biophysica Acta (BBA) - Molecular Basis of Disease*, Molecular Genetics of Human Reproductive Failure 1822:1921–1930. doi:10.1016/j.bbadis.2012.06.013

McCoy RC. 2017. Mosaicism in preimplantation human embryos: when chromosomal abnormalities are the norm. Trends Genet 33:448–463. doi:10.1016/j.tig.2017.04.001

Munné S, Kaplan B, Frattarelli JL, Child T, Nakhuda G, Shamma FN, Silverberg K, Kalista T, Handyside AH, Katz-Jaffe M, Wells D, Gordon T, Stock-Myer S, Willman S, Acacio B, Lavery S, Carby A, Boostanfar R, Forman R, Sedler M, Jackson A, Jordan K, Schoolcraft W, Katz-Jaffe M, McReynolds S, Schnell V, Loy R, Chantilis S, Ku L, Kaplan B, Frattarelli J, Morales A, Craig HR, Perloe M, Witz C, Wang W-H, Wilcox J, Norian J, Thompson SM, Chen S, Garrisi J, Walmsley R, Mendola R, Shamma FN, Pang S, Sakkas D, Rooney K, Sneeringer R, Glassner M, Stock-Myer S, Wilton L, Martic M, Coleman P, Shepley S, Nakhuda G, Child T, Mounce G, Griffiths T, Feinberg RF, Blauer K, Reggio B, Rhinehart R, Ziegler W, Ahmed H, Kratka S, Willman S, Rosenbluth E, Ivani K, Thyer A, Silverberg K, Minter T, Miller C, Gysler M, Saunders P, Casper R, Conway D, Gordon T, Hughes M, Large M, Blazek J, Munné S, Wells D, Fragouli E, Alfarawati S. 2019. Preimplantation genetic testing for aneuploidy versus morphology as selection criteria for single frozen-thawed embryo transfer in good-prognosis patients: a multicenter randomized clinical trial. Fertility and Sterility 112:1071–1079.e7. doi:10.1016/j.fertnstert.2019.07.1346

Munné S, Spinella F, Grifo J, Zhang J, Beltran MP, Fragouli E, Fiorentino F. 2020. Clinical outcomes after the transfer of blastocysts characterized as mosaic by high resolution Next Generation Sequencing-further insights. European Journal of Medical Genetics 63:103741. doi:10.1016/j.ejmg.2019.103741

Navratil R, Horak J, Hornak M, Kubicek D, Balcova M, Tauwinklova G, Travnik P, Vesela K. 2020. Concordance of various chromosomal errors among different parts of the embryo and the value of re-biopsy in embryos with segmental aneuploidies. Molecular Human Reproduction 26:269–276. doi:10.1093/molehr/gaaa012

Popovic M, Dhaenens L, Taelman J, Dheedene A, Bialecka M, De Sutter P, Chuva de Sousa Lopes SM, Menten B, Heindryckx B. 2019. Extended in vitro culture of human embryos demonstrates the complex nature of diagnosing chromosomal mosaicism from a single trophectoderm biopsy. Human Reproduction 34:758–769. doi:10.1093/humrep/dez012

R Core Team. 2022. R: A Language and Environment for Statistical Computing.

Ren Y, Yan Z, Yang M, Keller L, Zhu X, Lian Y, Liu Q, Li R, Zhai F, Nie Y, Yan L, Smith GD, Qiao J. 2022. Regional and developmental characteristics of human embryo mosaicism revealed by single cell sequencing. PLOS Genetics 18:e1010310. doi:10.1371/journal.pgen.1010310

Roberts SA, Wilkinson J, Vail A, Brison DR. 2022. Does PGT-A improve assisted reproduction treatment success rates: what can the UK Register data tell us? J Assist Reprod Genet 39:2547–2554. doi:10.1007/s10815-022-02612-y

Sanders KD, Silvestri G, Gordon T, Griffin DK. 2021. Analysis of IVF live birth outcomes with and without preimplantation genetic testing for aneuploidy (PGT-A): UK Human Fertilisation and Embryology Authority data collection 2016–2018. J Assist Reprod Genet 38:3277–3285. doi:10.1007/s10815-021-02349-0

Scott RT, Upham KM, Forman EJ, Zhao T, Treff NR. 2013. Cleavage-stage biopsy significantly impairs human embryonic implantation potential while blastocyst biopsy does not: a randomized and paired clinical trial. Fertility and Sterility 100:624–630. doi:10.1016/j.fertnstert.2013.04.039

Swinbank R, Purser J. 2006. Fibonacci grids: A novel approach to global modelling. Quarterly Journal of the Royal Meteorological Society 132:1769–1793. doi:10.1256/qj.05.227

Tiegs AW, Tao X, Zhan Y, Whitehead C, Kim J, Hanson B, Osman E, Kim TJ, Patounakis G, Gutmann J, Castelbaum A, Seli E, Jalas C, Scott RT. 2021. A multicenter, prospective, blinded, nonselection study evaluating the predictive value of an aneuploid diagnosis using a targeted next-generation sequencing–based preimplantation genetic testing for aneuploidy assay and impact of biopsy. Fertility and Sterility 115:627–637. doi:10.1016/j.fertnstert.2020.07.052

Tutt DAR, Silvestri G, Serrano-Albal M, Simmons RJ, Kwong WY, Guven-Ates G, Canedo-Ribeiro C, Labrecque R, Blondin P, Handyside AH, Griffin DK, Sinclair KD. 2021. Analysis of bovine blastocysts indicates ovarian stimulation does not induce chromosome errors, nor discordance between inner-cell mass and trophectoderm lineages. Theriogenology 161:108–119. doi:10.1016/j.theriogenology.2020.11.021

Victor A, Ogur C, Thornhill A, Griffin DK. 2020. Preimplantation Genetic Testing for Aneuploidies: Where We Are and Where We’re Going 25–48. doi:10.1201/9780429445972-3

Victor AR, Griffin DK, Brake AJ, Tyndall JC, Murphy AE, Lepkowsky LT, Lal A, Zouves CG, Barnes FL, McCoy RC, Viotti M. 2019. Assessment of aneuploidy concordance between clinical trophectoderm biopsy and blastocyst. Human Reproduction 34:181–192. doi:10.1093/humrep/dey327

Viotti M. 2020. Preimplantation Genetic Testing for Chromosomal Abnormalities: Aneuploidy, Mosaicism, and Structural Rearrangements. Genes 11:602. doi:10.3390/genes11060602

Viotti M, Greco E, Grifo JA, Madjunkov M, Librach C, Cetinkaya M, Kahraman S, Yakovlev P, Kornilov N, Corti L, Biricik A, Cheng E-H, Su C-Y, Lee M-S, Bonifacio MD, Cooper AR, Griffin DK, Tran DY, Kaur P, Barnes FL, Zouves CG, Victor AR, Besser AG, Madjunkova S, Spinella F. 2023. Chromosomal, Gestational, and Neonatal Outcomes of Embryos Classified as Mosaic by Preimplantation Genetic Testing for Aneuploidy. Fertility and Sterility. doi:10.1016/j.fertnstert.2023.07.022

Viotti M, Victor AR, Barnes FL, Zouves CG, Besser AG, Grifo JA, Cheng E-H, Lee M-S, Horcajadas JA, Corti L, Fiorentino F, Spinella F, Minasi MG, Greco E, Munné S. 2021. Using outcome data from one thousand mosaic embryo transfers to formulate an embryo ranking system for clinical use. Fertil Steril 115:1212–1224. doi:10.1016/j.fertnstert.2020.11.041

Wang L, Wang X, Liu Y, Ou X, Li M, Chen L, Shao X, Quan S, Duan J, He W, Shen H, Sun L, Yu Y, Cram DS, Leigh D, Yao Y. 2021. IVF embryo choices and pregnancy outcomes. Prenatal Diagnosis 41:1709–1717. doi:10.1002/pd.6042

Wickham H. 2016. ggplot2: Elegant Graphics for Data Analysis. New York: Springer-Verlag.

Yan J, Qin Y, Zhao H, Sun Y, Gong F, Li R, Sun X, Ling X, Li H, Hao C, Tan J, Yang J, Zhu Y, Liu F, Chen D, Wei D, Lu J, Ni T, Zhou W, Wu K, Gao Y, Shi Y, Lu Y, Zhang T, Wu W, Ma X, Ma H, Fu J, Zhang J, Meng Q, Zhang H, Legro RS, Chen Z-J. 2021. Live Birth with or without Preimplantation Genetic Testing for Aneuploidy. N Engl J Med 385:2047–2058. doi:10.1056/NEJMoa2103613

Yang M, Rito T, Metzger J, Naftaly J, Soman R, Hu J, Albertini DF, Barad DH, Brivanlou AH, Gleicher N. 2021. Depletion of aneuploid cells in human embryos and gastruloids. Nat Cell Biol 23:314–321. doi:10.1038/s41556-021-00660-7

